# Effectiveness of Localized Lockdowns in the COVID-19 Pandemic

**DOI:** 10.1101/2020.08.25.20182071

**Authors:** Yige Li, Eduardo A. Undurraga, José R. Zubizarreta

**Affiliations:** Department of Health Care Policy, Harvard Medical School, 180A Longwood Avenue, Office 307-Z, Boston, MA 02115; Department of Biostatistics, Harvard School of Public Health, Huntington Avenue, Boston, MA 02115, USA; Escuela de Gobierno, Pontificia Universidad Católica de Chile, Vicuña Mackenna 4860, Macul, Santiago 7820436, Chile; Millennium Initiative for Collaborative Research in Bacterial Resistance (MICROB-R), Chile; Research Center for Integrated Disaster Risk Management (CIGIDEN), Santiago, Region Metropolitana, Chile; CIFAR Azrieli Global Scholars program, CIFAR, Toronto, Canada; Department of Statistics, Harvard Faculty of Arts and Sciences, 1 Oxford Street, Cambridge, MA 02138, USA

**Keywords:** Causal Inference, COVID-19, Localized Lockdowns, Non-Pharmaceutical Interventions

## Abstract

Non-pharmaceutical interventions, such as social distancing and lockdowns, have been essential to control the COVID-19 pandemic. In particular, localized lockdowns in small geographic areas have become an important policy intervention to prevent viral spread in cases of resurgence. These localized lockdowns can result in lower social and economic costs compared to larger-scale suppression strategies. Using an integrated dataset from Chile (March 3 through June 15, 2020) and a novel synthetic control approach, in this paper we estimate the effect of localized lockdowns, disentangling its direct and indirect causal effects on SARS-CoV-2 transmission. Our results show that the effects of localized lockdowns are strongly modulated by their duration and are influenced by indirect effects from neighboring geographic areas. Our estimates suggest that extending localized lockdowns can slow down the pandemic; however, localized lockdowns on their own are insufficient to control pandemic growth in the presence of indirect effects from contiguous neighboring areas that do not have lockdowns. These results provide critical empirical evidence about the effectiveness of localized lockdowns in interconnected geographic areas.

---

Since the beginning of the COVID-19 pandemic, non-pharmaceutical interventions have been essential to control and prevent the transmission of the severe acute respiratory syndrome-coronavirus 2 (SARS-CoV-2) (1-3). Non-pharmaceutical interventions range from simple individual-level recommendations, such as wearing face masks, frequent hand-washing, or maintaining physical distance, to society-level regulatory actions, such as school closures, quarantines, or lockdowns (1, 3). Efforts to control pandemic growth based on these interventions have been successful in some countries (4-7). The effects of non-pharmaceutical interventions have been described primarily using compartmental models (1, 3, 6, 8, 9), with the results informing policies around the world since the start of the pandemic (10, 11). In this paper, we adopt a complementary approach from the causal inference literature (12, 13). Arguably, health policy impact evaluations require a variety of study designs, data sets, and analytic approaches that support each other to provide stronger evidence (14). In general terms, this approach seeks to estimate the causal effect of localized lockdowns by approximating the hypothetical randomized experiment (trial) that would have been conducted under ideal circumstances to evaluate the policy in question (15-18).

As the COVID-19 pandemic develops across countries, policymakers need evidence to help them decide when and how to ease mobility restrictions or strengthen these restrictions in cases of resurgence. Even now that countries have started vaccinating, large-scale non-pharmaceutical interventions continue to be important, particularly in low- and middle-income countries, to avoid large increases in the number of cases (19). In this context, localized lockdowns have become an increasingly relevant policy option (20-25).

Localized lockdowns are typically implemented in transmission hotspots and can be applied to populations or areas large and small to suppress an outbreak. Localized lockdowns had not been widely used as a public health response to contain outbreaks until the current pandemic (20-25). In principle, localized lockdowns impose fewer social and economic costs compared to larger-scale SARS-CoV-2 suppression strategies and are thus more sustainable. They can also provide a gradual exit from nationwide lockdowns. Early in the pandemic, for example, the Chinese government imposed a localized lockdown and other strict non-pharmaceutical interventions in the city of Wuhan (6), effectively suppressing SARS-CoV-2 transmission (26). Subsequently, governments have implemented localized lockdowns in neighborhoods (e.g., Beijing, China), suburbs (e.g., Melbourne, Australia), towns (e.g., Vo, Italy), districts (e.g., North Rhine-Westphalia, Germany), and at the city level in Leicester, England (4, 21). Despite the increasing importance of localized lockdowns, there is limited empirical evidence of their effectiveness.

In this study, we use data from Chile to estimate the effect of localized lockdowns on COVID-19 transmission. Our data set combines information from administrative COVID-19 surveillance records (27), a nationally representative household survey (28), and census data (29). We use a synthetic control approach (30, 31) to build control intervention units (municipalities) with similar sociodemographic features, trajectories of contagion, and histories of lockdowns until the time of the policy intervention, taking into account the spatio-temporal structure of the data. In other words, we assess whether the effectiveness of a localized lockdown implemented at the municipality level was affected by the lockdown status of its neighboring municipalities. This indirect effect may play an important role in municipalities within cities or urban areas where social and economic interdependencies exist. Allowing for such indirect effects or interference between municipalities (32, 33), we estimate the direct effects of extending the duration of localized lockdowns and the total (sum of direct and indirect) effects of maintaining lockdowns in neighboring municipalities.

## METHODS

### Lockdowns in Chile

The Ministry of Health reported the first COVID-19 case in Chile on March 3, 2020 (34). By the end of March, the government had restricted large gatherings (March 13), closed schools and universities (March 16), increased controls on national borders (March 18), enforced night-time curfews (March 22), and imposed mandatory use of facemasks in public (April 8) (34) (Figure 1). These policies and recommendations were implemented uniformly nationwide, with no within-country variation until July 19, 2020 (34). During the study period (March 3 through June 15, 2020), the only exceptions were localized lockdowns, implemented at the municipal level, the smallest administrative subdivision in Chile. Starting on July 19, 2020, the government initiated a gradual reopening scheme of five incremental steps, also implemented at the municipal level (35).

**Figure 1.**
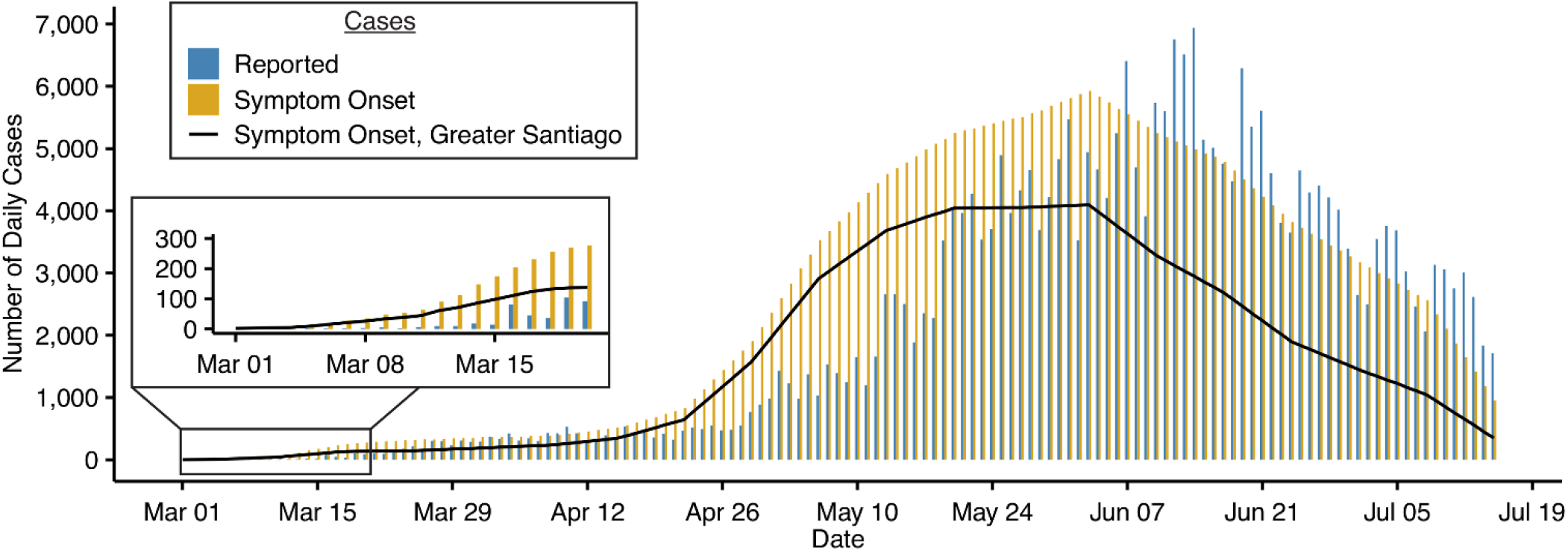
Incidence of COVID-19 cases in Chile, March 1 to July 15, 2020. The majority (66%) of cases were reported in Greater Santiago (solid line).

In Chile, localized lockdowns were implemented at various points in time, typically at the municipality level, although in some cases they involved only a portion of its population (Figure 2A, 2B). The government loosely defined the criteria used to impose lockdowns as a function of the number and density (per km^2^) of infectious COVID-19 cases, increases in case incidence, and health system capacity (34). Across the country, there was substantial variation in the duration of these municipal-level localized lockdowns and, for each municipality under lockdown, in the lockdown status of neighboring municipalities. As a result, the effectiveness of lockdowns varied geographically. We used this policy variation as a natural experiment to evaluate the effectiveness of localized lockdowns on SARS-CoV-2 transmission.

**Figure 2.**
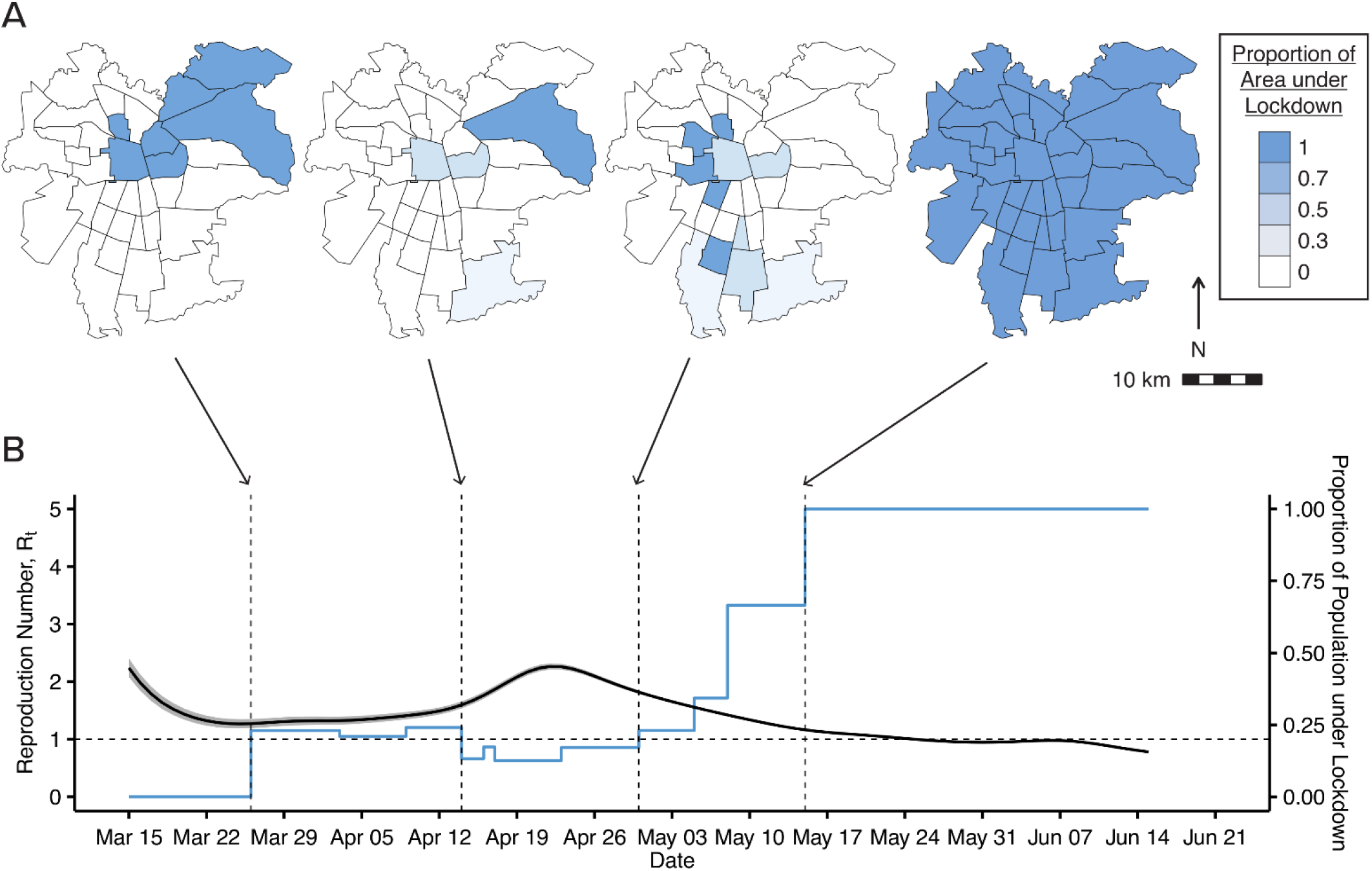
Lockdowns and instantaneous reproduction number. (A) Municipality-level lockdowns in Greater Santiago on March 31, April 15, April 30, and May 15, 2020. (B) Variation of the instantaneous reproduction number (*R*_*t*_) and the proportion of the population under lockdown in Greater Santiago over time.

### Study overview

Based on the potential outcomes framework for causal inference (12, 13, 36, 37), we used the augmented synthetic control method to analyze the pandemic’s progression in comparable municipalities that underwent different lockdown interventions (30, 31). For an individual municipality, we varied the duration of the intervention and the proportion of the population under lockdown in the neighboring municipalities at each time point, controlling for relevant covariates that could confound these effects (Figure 3B). We estimated the counterfactual progression that the disease would have exhibited had an alternative lockdown policy taken place in the focal municipality or its neighbors (Figure 3B). We provide open-source code with step-by-step explanations to replicate the analyses and implement them in related settings (see the Supplementary Material).

**Figure 3.**
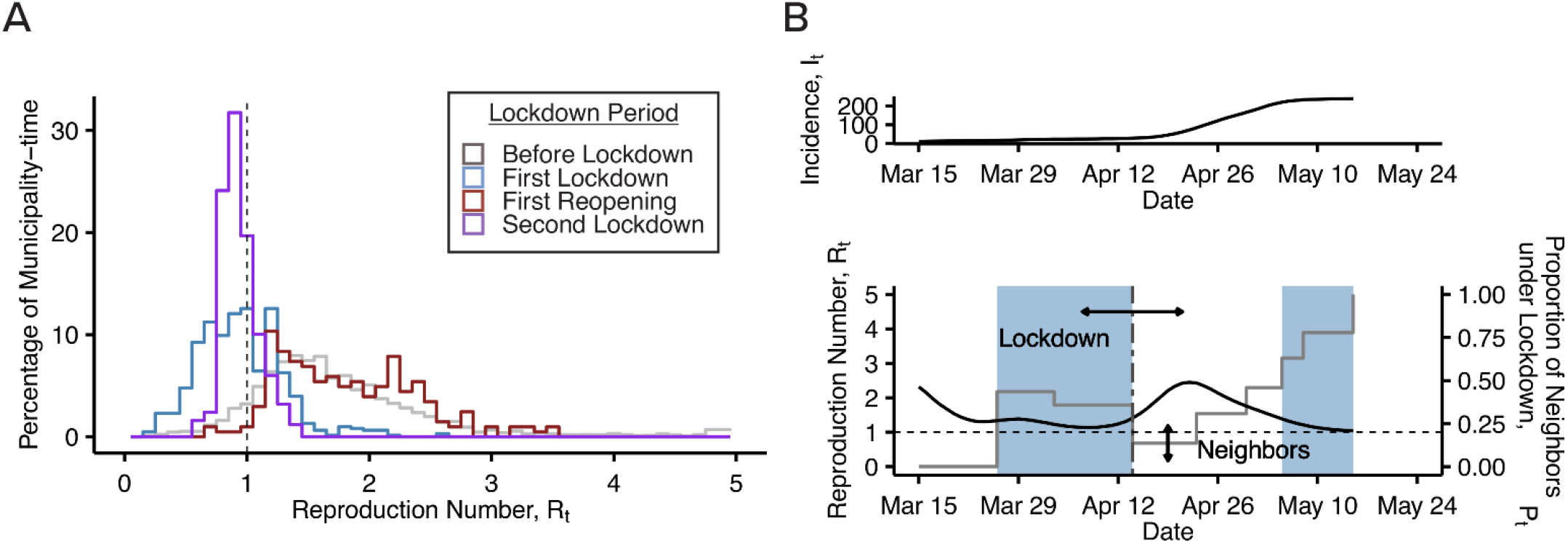
Histogram of daily *R*_*t*_ and alternative lockdown interventions. (A) Histogram of daily *R*_*t*_ for all municipalities in Chile from March 15 to June 15, 2020, divided into the periods before lockdown, during the first lockdown, during the first reopening, and during the second lockdown. Municipalities with fewer than 10 COVID-19 cases were excluded. (B) Illustration of alternative lockdown interventions of varying durations (blue area) and proportion of neighbors under lockdown (grey line).

### Integrated surveillance records, survey measurements, and census data

Our data set combines information from administrative COVID-19 surveillance records, a nationally representative household survey, and census data. Specifically, we use epidemiologic surveillance records from the Department of Epidemiology of the Chilean Ministry of Health (34). COVID-19 cases are defined in our dataset as symptomatic and asymptomatic SARS-CoV-2 infections confirmed by a positive PCR test. Throughout the pandemic, Chile has tested for COVID-19 at a higher rate than any other Latin American country (38, 39) and features a high level of effective universal health coverage (40). We characterized municipalities based on Chile’s National Socioeconomic Characterization Survey (CASEN), a nationally representative household survey that collects data on education, employment, income, health, and housing (28). Finally, we employed population data from the 2017 National Census (29). All data are publicly available.

We adjusted the COVID-19 case incidence series to correct the lag in reporting and some incomplete municipality-level data. First, we imputed the incomplete data by interpolating between the closest dates with complete data. In the data, the number of cumulative cases was reported typically every 2-4 days. Second, we estimated the lag in reporting using the PELT algorithm (41). Third, we adjusted the incidence series for space- and time-varying reporting lags using the approach by Zhao et al. (42). These adjustments considered that the lag between symptom onset date and report date could vary across municipalities and over time. We estimated the instantaneous reproduction number following Cori et al. (43) using the adjusted COVID-19 series. See the Supplementary Material for details.

### Instantaneous reproduction number

We characterized transmission by the instantaneous reproduction number (*R*_*t*_); that is, the average number of secondary cases per primary infected case (43). We did this using the daily series of COVID-19 cases reported by the Ministry of Health (34), adjusted for the time-lag between onset of symptoms and case report (Figure 1) (42, 43). Following Cori et al. (43), the instantaneous reproduction number can be estimated by 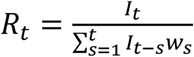, where *I*_*t*_ is the incidence at time *t* and *w*_*s*_ is the infectivity function or density of the serial interval at time *s*. Cori et al. (43) propose estimating *R*_*t*_ over a window of time *τ* as

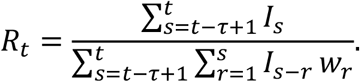

See Gostic et al.(44) for a discussion of the method by Cori et al.(43) and other related approaches to estimating the instantaneous reproduction number. Based on data from the SARS-CoV-2 outbreak in Wuhan, Li et al. (45) estimated that the serial interval distribution had a mean of 7.5 days, a standard deviation of 3.4 days, and a 95% confidence interval of [5.3, 19] days. Thus, following Cori et al. (43), we assumed that the serial interval has a Gamma distribution with a mean of 7.5 days and a standard deviation of 3.4 days and *τ* = 5. Assuming homogeneous mixing of the population within a municipality, we calculated *R*_*it*_ for each municipality *i* (Figure 3A).

### Potential outcomes

We base our analysis on the potential outcomes framework for causal inference (13, 36, 37) under interference (32, 33, 46). Let 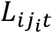 denote the lockdown indicator for municipality *i =* 1, *…, I* in cluster *j*_*i*_ *=* 1, *…, J*_*i*_ at time *t =* 1, *…, T*, with 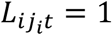 if the municipality is under lockdown at time *t*, and 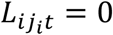 otherwise. In our analysis, since the cluster of the municipality *i* is defined as the union of *i* and its adjacent municipalities, we can omit the index *j*_*i*_ for simplicity in the notation, but our approach is more general. Write *<u>L</u>*_*it*_ for the lockdown history of municipality *i* until time *t*. Analogously, define *P*_*(i)t*_ as the proportion of the population in the cluster of municipality *i* under lockdown at time *t*, excluding municipality *i*, and *<u>P</u>*_*(i)t*_ as the corresponding history until time *t*. In this framework, *L*_*it*_ can also denote the proportion of the population under lockdown in municipality *i*, beyond whether or not the municipality is under lockdown. Following the frameworks for causal inference under interference by Sobel (32) and Hudgens and Halloran (33), we put *R*_*it*_ (*<u>L</u>*_*it*_ *= <u>l</u>*_*it*_, *<u>P</u>*_*(i)t*_ *= <u>p</u>*_*(i)t*_) for the potential instantaneous reproduction number for municipality *i* in its cluster at time *t* under lockdown histories *<u>l</u>*_*it*_ and *<u>p</u>*_*(i)t*_ for the municipality and its neighbors until time *t*, respectively. Finally, we designate *I*_*it*_ (*<u>I</u>*_*it−*1_(*), *R*_*it*_ (*<u>L</u>*_*it*_ *= <u>l</u>*_*it*_, *<u>P</u>*_*(i)t*_ *= <u>p</u>*_*(i)t*_)) as the potential incidence in municipality *i* at time *t*, which is a function of its potential incidence history *I*_*it−*1_(*) and potential instantaneous reproduction number *R*_*it*_ (*<u>L</u>*_*it*_ *= <u>l</u>*_*it*_, *<u>P</u>*_*(i)t*_ *= <u>p</u>*_*(i)t*_).

### Direct, indirect, and total effects of lockdowns

For any given municipality *i*, we aim to estimate the effect of lockdowns on the instantaneous reproduction number *R*_*it*_ when intervening both on the duration of the lockdown *<u>L</u>*_*it*_ and on the proportion of the population under lockdown in the neighboring municipalities *<u>P</u>*_*(i)t*_. We are particularly interested in the municipality-level direct, indirect, and total effects of lockdowns across time (33). See the Supplementary Material for precise definitions of these estimands.

### Synthetic controls

We used the augmented synthetic control method (30, 31) to estimate the potential instantaneous reproduction number *R*_*it*_(*) and calculate the potential incidence *I*_*it*_(*) from *R*_*it*_(*) according to 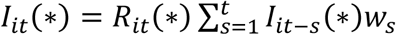 (see the Supplementary Material for details). The intuition behind this method is, for a given lockdown intervention in municipality *i*, to build a synthetic control municipality with very similar covariate, intervention, and outcome histories leading up to the time of the intervention by appropriately weighting control municipalities over time. We adjusted for (or balanced) several municipality-level characteristics that may affect virus transmission (see Table S2 in the Supplementary Material). Adjustments included the proportion of females in the population, older than 65 years of age, in rural areas, under poverty, in overcrowded households (≥2.5 people per room), with inadequate sanitation infrastructure (access to potable water and sewage), average monthly income, and municipality area.

Furthermore, we adjusted for the seven-day history of lockdown interventions in the municipality *<u>L</u>*_*i*[*t*−7,*t*−1]_and its neighbors *<u>P</u>*_*i*[*t*−7,*t*−1]_, and for the instantaneous reproduction number *<u>R</u>*_*i*[*t*−7,*t*−1]_. We analyzed all the municipalities in Greater Santiago that started their first lockdown after March 15, 2020, and completed it by May 15, 2020 (Figure 2A); that is, the first period of confinement that arguably shaped the evolution of the pandemic (Figure 1, Figure 2B).

## RESULTS

### Balance and predictive power

All intervention and corresponding synthetic control municipalities were closely balanced in terms of their baseline sociodemographic characteristics *X*_*i*_, in addition to their seven-day trajectories *<u>L</u>*_*i*[*t*−7,*t*−1]_, *<u>P</u>*_*i*[*t*−7,*t*−1]_, and *<u>R</u>*_*i*[*t*−7,*t*−1]_(see tables S1-S6 in the Supplementary Material). In terms of predictive power, the out-of-sample proportion of the variation of the outcomes explained by the synthetic controls (defined by 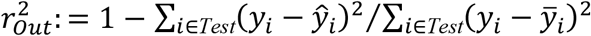, where *y* denotes the outcome and *Test* comprises its out-of-sample observations in the post-intervention period, which extended up to 21 days after the intervention) was 64% for the instantaneous reproduction number *R*_*t*_ and 92% for the incidence *I*_*t*_.

### Duration and indirect effects of localized lockdowns

Overall, our results suggest that the effectiveness of localized lockdowns is strongly modulated by the duration of the intervention and the magnitude of the indirect effects from neighboring geographic areas. The larger the proportion of neighbors under lockdown, the higher the effectiveness of the lockdown in controlling transmission.

We illustrate our findings with three representative municipalities in Greater Santiago that were placed under lockdown on March 26: Lo Barnechea, Providencia, and Santiago (Figure 4; see the Supplementary Material for results from other municipalities). We chose these municipalities because they were among the first to be placed under lockdown, and they have substantial interdependence with other municipalities in Greater Santiago. The municipality of Santiago concentrates the country’s financial, commercial, and political activity, including all major government infrastructure. Providencia is an upper-middle-class urban municipality with substantial commercial activity, high-rise apartment buildings, and the highest proportion of the population over 65 years of age. Lo Barnechea is primarily a residential area, with limited commercial activity, few buildings, and a heterogeneous population. In the Supplementary Material (Figures S3-S6) we present additional results for other municipalities in Greater Santiago (Ñuñoa, Independencia, Las Condes, Vitacura) and elsewhere in Chile (Arica and Punta Arenas, the northernmost and southernmost cities in Chile, respectively); the findings for these municipalities are consistent with our main results.

**Figure 4.**
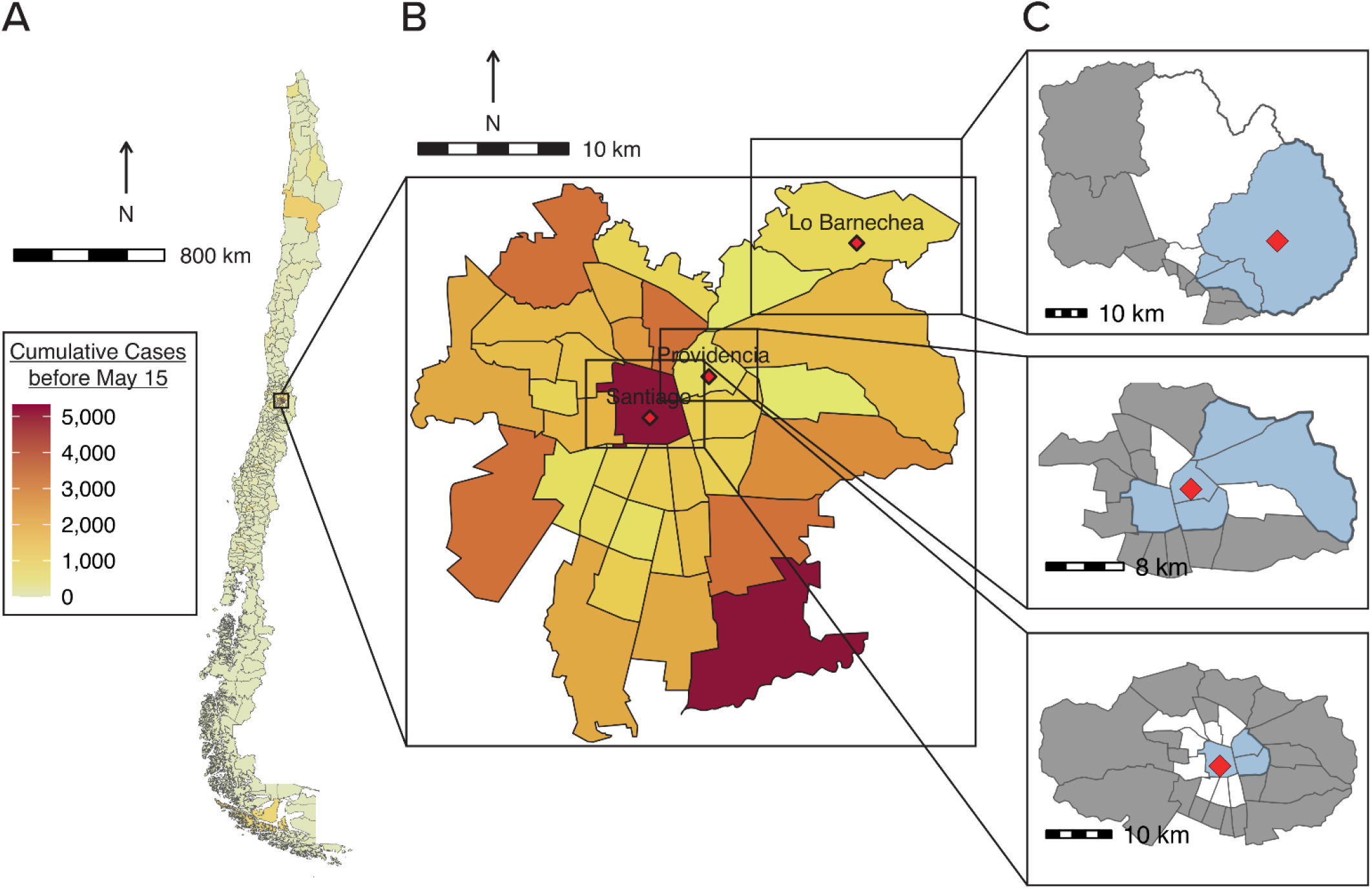
Distribution of cumulative cases of COVID-19 and illustration of lockdown implementation. (A) Cumulative COVID-19 cases in Chile and (B) Greater Santiago before May 15, 2020. (C) Municipalities of Lo Barnechea, Providencia, and Santiago (red diamonds), and their corresponding neighbors (blue for the immediate neighbors under lockdown and white otherwise) on the last date under lockdown, April 13.

Figure 5 shows a large reduction in *R*_*t*_ (Fig. 5A) and COVID-19 cases (Figure 5B) with an extended lockdown. Had the lockdown been extended for three additional weeks, maintaining *P*_*t*_ constant, we estimate that the reduction in *R*_*t*_ would have been larger. The average *R*_*t*_ would have decreased from 1.83 to 1.27 (difference: −0.56, 95% confidence interval [CI]: [-0.63,-0.50]) in Lo Barnechea, from 1.82 to 1.34 (difference: −0.47, 95%CI: [-0.59,-0.36]) in Providencia, and from 1.95 to 1.23 (difference: −0.72, 95%CI: [-0.85,-0.58]) in Santiago. These reductions in *R*_*t*_ are equivalent to 177 (95%CI: [167,188]; or 143 per 100,000 population) averted COVID-19 cases over three weeks in Lo Barnechea, 94 (95%CI: [76,111]; or 59 per 100,000 population) averted cases in Providencia, and 1343 (95%CI: [1245,1441]; 267 per 100,000 population) averted cases in Santiago, which would represent 33-62% reductions in reported cases in that time frame.

**Figure 5.**
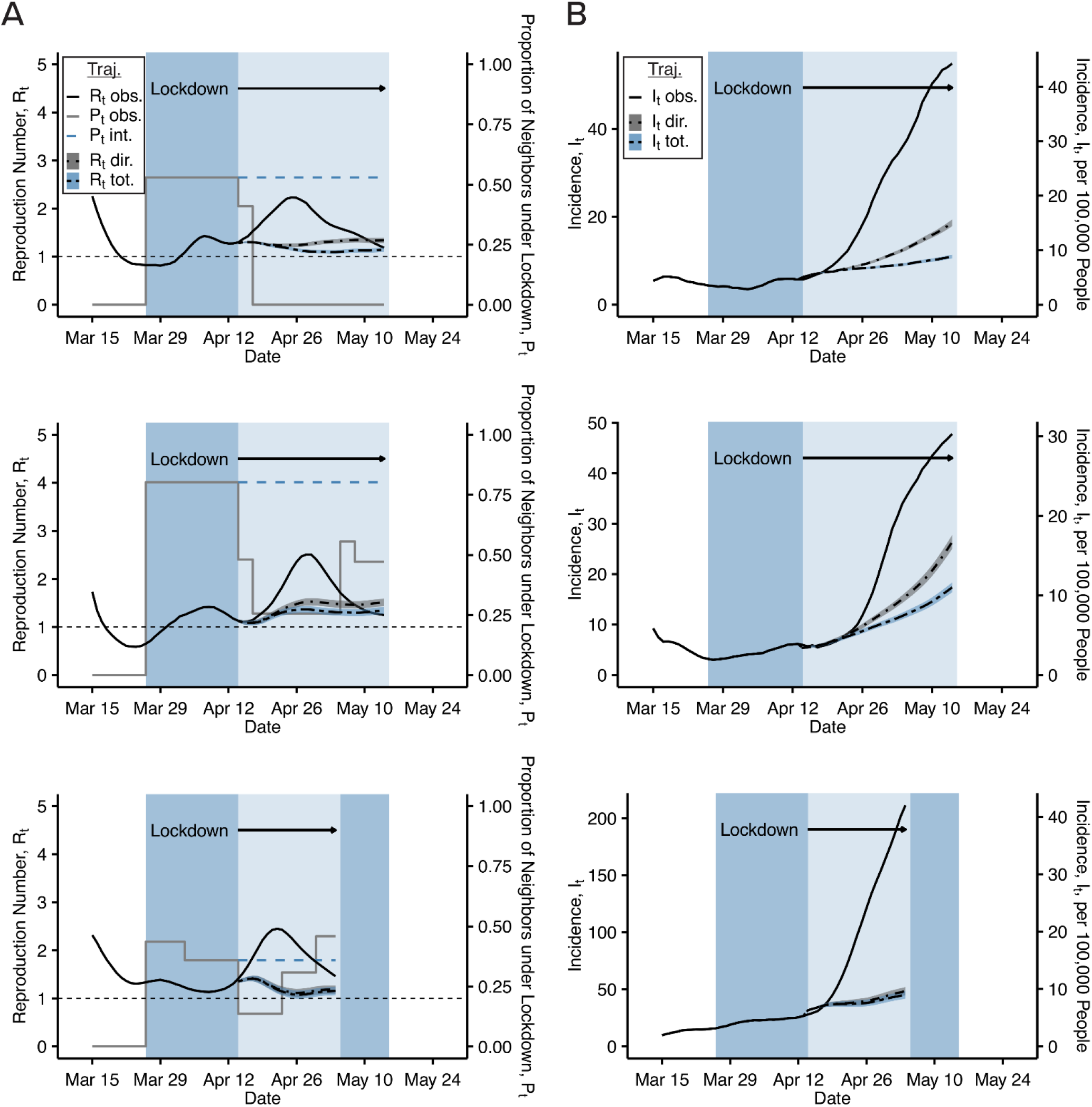
Estimated instantaneous reproduction number *R*_*t*_ and incidence *I*_*t*_ under different lockdown interventions by municipality. In (A), the grey solid and blue dashed lines show the proportion of the population in the surrounding municipalities under lockdown, respectively for the observed and the intervened *P*_*t*_ (*P*_*t*_ *obs* and *P*_*t*_ *int*). In (A, B), the lighter shade of blue extends the duration of the lockdown. The solid black lines show the reproduction number *R*_*t*_ (A) and daily incidence *I*_*t*_ (B), and the dashed lines show the predicted *R*_*t*_ and case incidence *I*_*t*_ for the extended lockdown. The direct and total effects of the extended lockdown are the difference between the solid black line and dashed line with grey (*R*_*t*_ *dir, I*_*t*_ *dir*) and blue (*R*_*t*_ *tot, I*_*t*_ *tot*) bands, respectively. The bands around the curves indicate 95% confidence intervals (additional results in the Supplementary Material).

The reductions in transmission would have been even larger if lockdowns in neighboring municipalities had taken place. Assuming municipalities adjacent to Lo Barnechea, Providencia, and Santiago maintained their lockdown status (*P*_*t*_ = 53.0%, *P*_*t*_ = 80.3%, and *P*_*t*_ = 35.8%) for three additional weeks, we estimate that the average *R*_*t*_ would have decreased to 1.19 (95%CI: 1.13, 1.25), 1.25 (95%CI: 1.14, 1.37), and 1.21 (95%CI: 1.08, 1.34), respectively (Figure 5A). Figures 6A and 6B show the relationship between daily COVID-19 incidence and days of extended lockdown as a function of changes in *P*_*t*_, after adjusting for observed covariates. The larger *P*_*t*_, the greater the number of averted cases. Overall, results in Greater Santiago suggest that the decision to reopen these municipalities was premature, especially when lockdowns were brief, because the effectiveness of lockdowns strongly depends on their duration and the magnitude of indirect effects (findings for other municipalities with lockdowns are consistent with these results; see Figures S3-S6 in the Supplementary Material).

**Figure 6.**
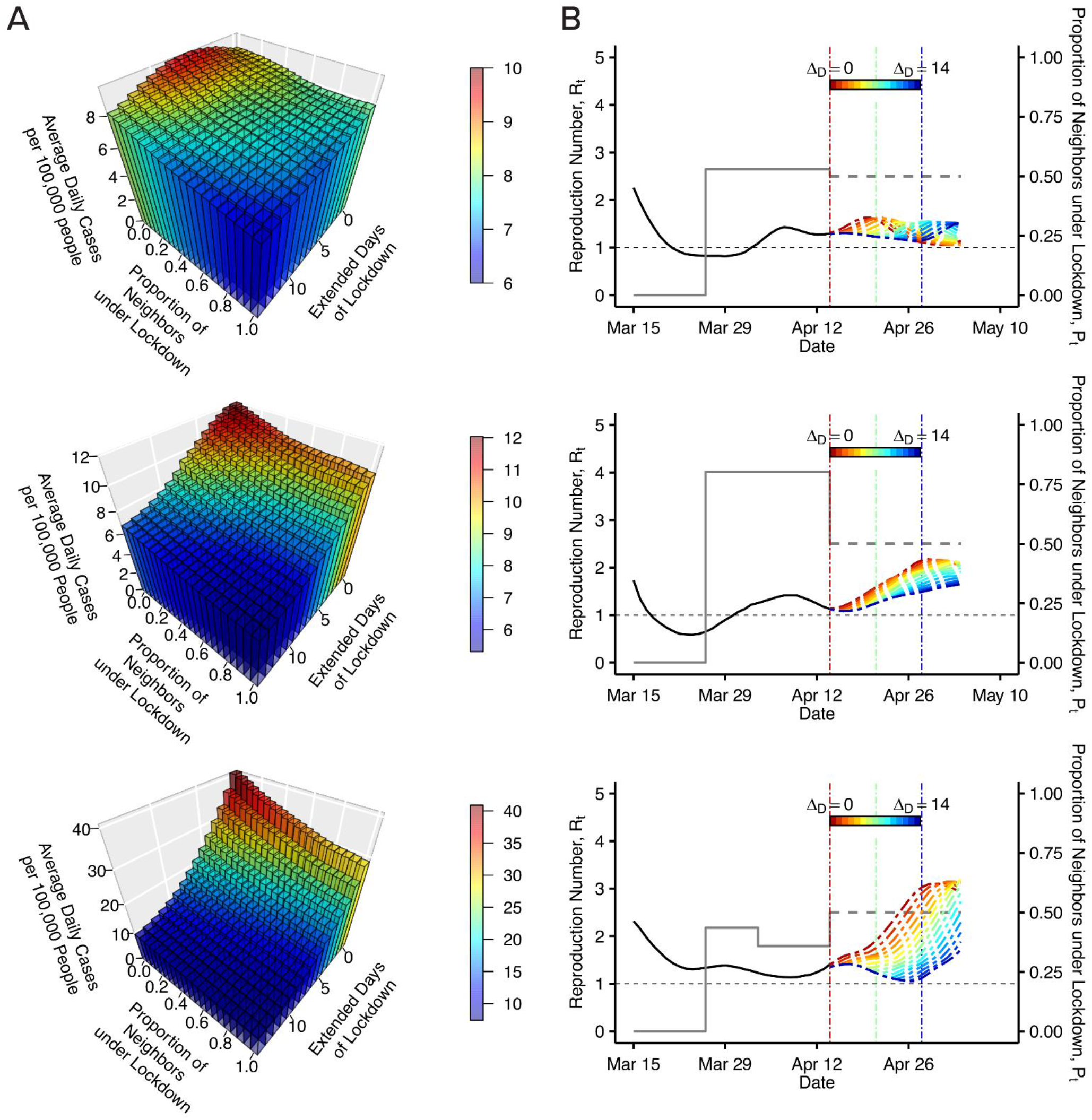
Duration and indirect effects strongly modulate the effectiveness of lockdowns. From top to bottom, the municipalities in the figures are Lo Barnechea, Providencia, and Santiago, March 15 to May 15, 2020. (A) Average daily COVID-19 incidence over three weeks with varying duration of extended lockdown (Δ_*D*_ = 0-14 days) and a varying proportion of the neighboring population under lockdown (*P*_*t*_ = 0-1). (B) Prediction of the instantaneous reproduction number *R*_*t*_ as a function of time with 50 percent of the neighboring population under lockdown (*P*_*t*_ = 0.5) since intervention (lockdown extended for 0-14 days).

### Lockdowns without indirect effects

Figure S5 in the Supplementary Material reaffirms the results from a different perspective. As happened with Lo Barnechea, Providencia, and Santiago, the municipality of Punta Arenas in the south of Chile was placed under lockdown early in the pandemic, from April 1 to May 7. It initiated lockdown with one of the highest case incidences per 100,000 population in the country. Notably, Punta Arenas is geographically isolated and has few local interdependencies that could result in active transmission networks during a localized lockdown. Our estimates show negligible indirect effects: increasing *P*_*t*_ from 0 to 1 would only result in a reduction of *R*_*t*_ of 0.02 (Figure S5 and Table S14), arguably due to its geographical isolation and minor interdependencies with neighboring municipalities.

### Varying the area under lockdown

Having assessed the role of duration and indirect effects, we evaluate the impact of lockdowns in geographic areas of increasing size. We considered three target lockdown areas (Figure 7A): the municipality of Ñuñoa (red), a cluster of six municipalities (orange), and Greater Santiago (green). We extended the study period to encompass the mandatory lockdown for Greater Santiago that began on May 15. We varied the population under lockdown in the targeted area and the proportion of the population under lockdown in neighboring municipalities (*P*_*t*_). Note that the municipality of Ñuñoa had only about half its population under lockdown between April 14 and May 7, as indicated by the blue line in Figure 7B. Figure 7B shows the estimated *R*_*t*_ from March 15 to June 15. In general, an epidemic will continue to grow as long as *R*_*t*_ is greater than one. Figure 7 shows that the pandemic kept expanding in all three target areas until a city-wide lockdown was implemented on May 15. These results highlight the challenges of suppressing virus transmission in areas with a high degree of economic and social interdependencies, such as Chile’s capital, where a substantial proportion of the municipalities were not under lockdown.

**Figure 7.**
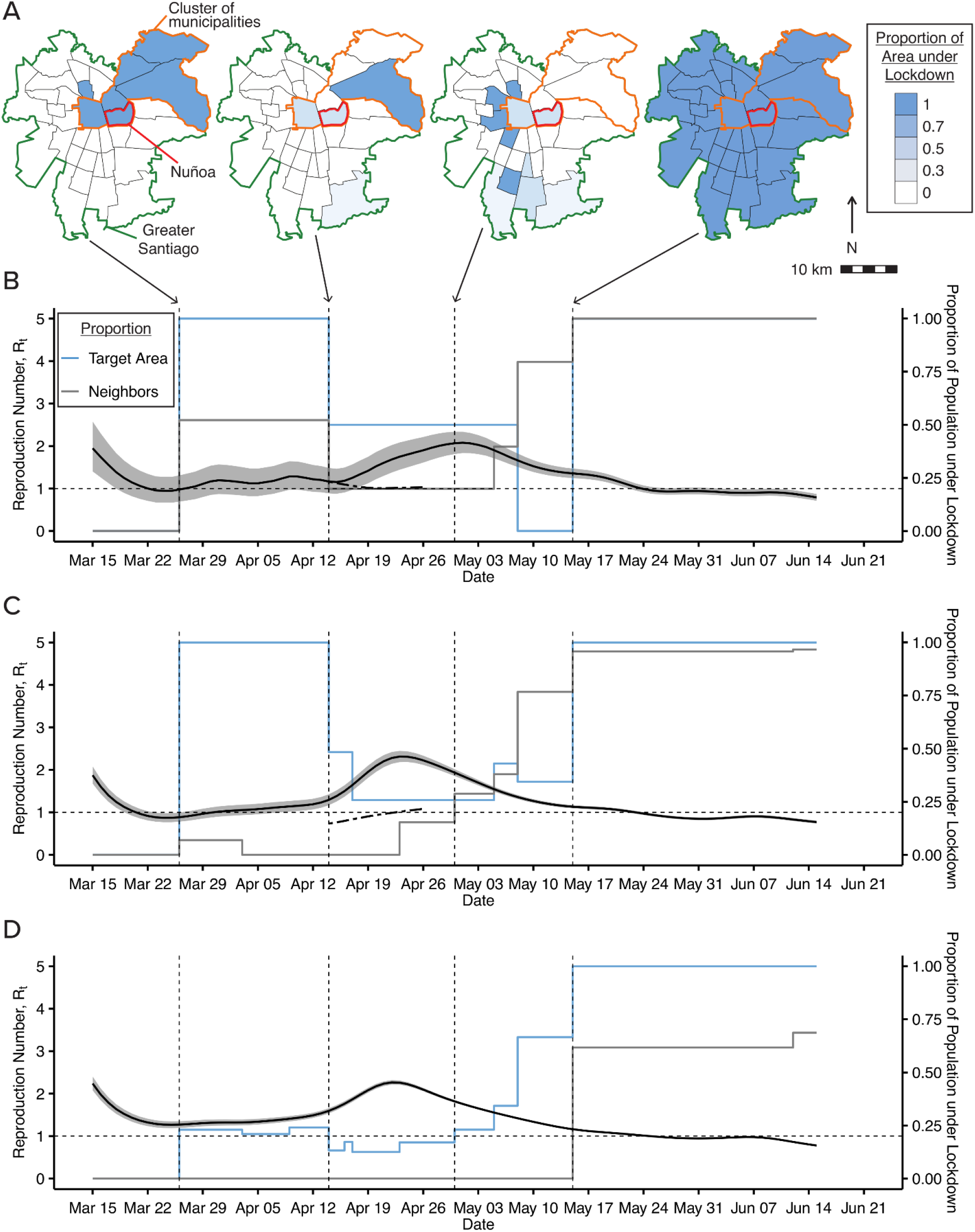
Effectiveness of lockdowns for different target areas. (A) In blue, municipalities of Greater Santiago under lockdown at different points in time; outlined in red (Ñuñoa), orange (cluster of six municipalities), and green (Greater Santiago), lockdown target areas of increasing size. (B-D) Estimated instantaneous reproduction number *R*_*t*_, with changing proportions of the population under lockdown in each geographic area and its immediate neighboring areas. From top to bottom the figure shows the municipality of Ñuñoa, a cluster of six municipalities, and the city Greater Santiago. (B-C) Also show the predicted *R*_*t*_ (dashed line) had the lockdowns in the geographic area and for its immediate neighbors been extended. We estimate that the epidemic would have continued to grow (*R*_*t*_ ≥ 1) even with the extended localized lockdowns. The epidemic kept growing until Greater Santiago was put under lockdown on May 15, 2020.

## DISCUSSION

Using augmented synthetic control methods, we estimated the effects of localized lockdowns on COVID-19 transmission, incorporating the effects of lockdown duration as well as lockdowns occurring in neighboring areas. We found that localized lockdowns can help contain the transmission of the virus. However, their effectiveness is dependent on lockdown duration and potential indirect effects from neighboring geographic areas with high social interconnectivity. For instance, the effectiveness of localized lockdowns within Greater Santiago, where there are high economic and social interdependencies between municipalities, was strongly affected by the extent of suppression measures concurrently in place in neighboring municipalities. As expected, as the proportion of neighbors under lockdown increased, so did the effectiveness of localized lockdowns in controlling pandemic spread. However, our estimates also show that in Greater Santiago, the epidemic is only controlled (i.e., *R*_*t*_ < 1) when generalized lockdowns are in place. In contrast, localized lockdowns showed promising results in municipalities such as Punta Arenas, which are geographically isolated and thus have transmission networks that are relatively unaffected by neighboring areas.

The marginal effectiveness of localized lockdowns may decline over time, as suggested by the results in Figure S7 (Supplementary Material). On one hand, lockdowns may be particularly challenging for socially vulnerable individuals who depend on daily wages, have limited savings, or do not receive external support, as their need to secure income may conflict with policy (47, 48). On the other hand, lockdowns may not immediately decrease *R*_*t*_ to their full potential because transmission within households or other group residences, such as nursing homes, account for a substantial proportion of new cases (6, 49). There are sustained close contacts within households, particularly where people cannot isolate themselves in a separate room or where members share common spaces such as restrooms (50-52). It is also possible that lockdowns may not be sufficient to contain an epidemic. For instance, *R*_*t*_ was significantly reduced in Wuhan after implementation of a city-wide lockdown. However, control of the pandemic (*R*_*t*_ < 1) was only achieved through a system of centralized quarantines and treatment for COVID-19 patients in field hospitals (6, 26). Our results suggest that the effectiveness of lockdowns depends upon their duration and on potential indirect effects from neighboring geographic areas; therefore, if interdependencies exist, then achieving *R*_*t*_ < 1 may not be possible.

To evaluate the effectiveness of localized lockdowns, our approach implicitly models the transmission process across municipalities by adjusting for the preceding seven-day trajectories of lockdown interventions in the municipality *<u>L</u>*_*i*[*t*−7,*t*−1]_, its neighbors *<u>P</u>*_*i*[*t*−7,*t*−1]_, and the instantaneous reproduction number *<u>R</u>*_*i*[*t*−7,*t*−1]_, in addition to the baseline sociodemographic characteristics *X*_*i*_. We assume that this process is similar for municipalities with comparable observed baseline characteristics and trajectories of lockdowns and contagion over time. In this context, our approach estimates the expected number of new cases (*R*_*t*_) from each infected case in the municipality, considering that some individuals may have infected elsewhere, and from this number calculates the incidence (*I*_*t*_) in the municipality. To reduce the complexity and increase the generalizability of our approach, we assumed that lockdowns affect only neighboring municipalities. We lack data on mobility between specific municipalities to test this hypothesis. In large urban areas, the degree of commuting beyond neighboring municipalities may be non-negligible. However, at least 50% of trips in Greater Santiago were walking, bicycle, or short-distance car trips (53), and mobility was significantly reduced during the pandemic. Estimates suggest traffic decreased by 54-59% between early March and the first week of June in Greater Santiago (54, 55). Also, lockdowns were enforced at the municipality level (34). When under lockdown, individuals need a valid police-issued permit for street travel. Our estimation approach can be modified to control for a network of influence that does not necessarily correspond to the nearest geographic neighbors but to the most influential entities through *<u>P</u>*_*(i)t*_ in the potential instantaneous reproduction number.

Our study centers on the first three months of the pandemic in Chile. During this period, the proportion of the susceptible population decreased slightly. While it is possible that changes in the susceptible population may affect the infection rates, these changes were limited during the study period. In fact, between March 3 and June 15, 2020, there were 179,436 reported COVID-19 cases and 4,883 deaths in a total population of nearly 19 million. As noted earlier, Chile has had the highest total testing rate in Latin America throughout the pandemic (56), and all cases are lab-confirmed by RT-PCR. Also, since our synthetic control approach adjusts for the 7-day moving history of *R*_*t*_, it captures the downward trend of *S*_*t*_*/N*, the ratio of the susceptible population at time *t* over the total population. In fact, for each intervention municipality at time *t*, when we move to build its synthetic control in time *t* + 1, we also move one period ahead in the control set. Therefore, the underlying susceptible proportions are decreasing in a similar way in the intervention municipality and its synthetic control.

Our analysis and the currently available data have limitations. First, we based our analysis on reported COVID-19 cases (27), which may be affected by underreporting. SARS-CoV-2 infection can result in a broad spectrum of clinical outcomes, including asymptomatic infection, mild symptoms, hospitalization, or death (57-59). Mild cases without apparent symptoms may be undetected (26, 60, 61). However, Chile has conducted tests at a substantially higher rate than any other country in Latin America, with approximately 130 total RT-PCR tests per 1,000 people by August 31, 2020 (39). Also, it is possible that differences in health-seeking behavior and the severity of illness, for example, by age group, may bias the COVID-19 case data. However, since the population characteristics hardly changed during the study period, this potential bias should be stable and not meaningfully affect the results as long as reporting rates are consistent in time. Second, in Chile the Ministry of Health does not report COVID-19 cases by the onset of symptoms but by the day of reporting to the health system. We addressed this limitation by adjusting the time series, according to Zhao et al. (42). Third, the causal validity of our estimates is predicated on identification assumptions (such as direct interference and ignorable treatment assignment) often invoked in related studies. While strong, these assumptions are necessary to identify the direct and indirect effects of lockdowns from the available data. See the Supplementary Material for further details.

It is well-known that the only way to stop an epidemic is to break the transmission chain. To this end, non-pharmaceutical interventions have been extensively used in the COVID-19 pandemic (62). Large-scale interventions have imposed high social and economic costs to societies worldwide (47, 63-65). As they are less disruptive than large-scale interventions, localized lockdowns can help reduce those costs and provide a more sustainable strategy over time (66). Also, localized lockdowns may provide a gradual, more controlled exit relative to larger-scale strategies if effectively implemented. In principle, localized lockdowns can break transmission chains by limiting contact between infectious and susceptible individuals, and this goal could be achieved at household, neighborhood, municipality, county, or state levels. However, the social distancing imposed by a lockdown must be maintained and enforced until adequate control of transmission is achieved. The effectiveness of non-pharmaceutical interventions depends on the willingness and capacity of the population to comply. Compliance may be particularly challenging in low- and middle-income countries, where a substantial proportion of the population works informally (67). Hence, the ability to comply with stay-at-home restrictions is at odds with the need to secure income on a daily basis (47, 68). While localized lockdowns may show promise in mitigating social costs and extending their length can be beneficial, this study shows that their effectiveness can be attenuated by indirect effects from neighboring areas where transmission networks exist, such as in cities. The growth of disease transmission is reversed only when lockdowns are implemented in a coordinated fashion across interconnected geographic areas.

## Supporting information

SuppMaterial_Li_etal_LocalLockdown

## Data Availability

Datasets used and/or analyzed during the current study are available from Base de Datos CoVID-19 repository, Ministerio de Ciencia, Tecnologia, Conocimiento, e Innovacion.

http://www.minciencia.gob.cl/covid19

## Author affiliations

Department of Biostatistics and CAUSALab, Harvard T.H. School of Public Health, Huntington Avenue, Boston, Massachusetts 02115 (Yige Li, José R. Zubizarreta); Escuela de Gobierno, Pontificia Universidad Católica de Chile, Vicuña Mackenna 4860, Macul, Santiago 7820436, Chile (Eduardo A. Undurraga); Millennium Initiative for Collaborative Research in Bacterial Resistance (MICROB-R), Chile (Eduardo A. Undurraga); CIFAR Azrieli Global Scholars program, CIFAR, Toronto, ON M5G 1M1, Canada (Eduardo A. Undurraga); Research Center for Integrated Disaster Risk Management (CIGIDEN), Santiago, Chile (Eduardo A. Undurraga); Department of Health Care Policy, Harvard Medical School, 180A Longwood Avenue, Office 307-Z, Boston, Massachusetts 02115 (José R. Zubizarreta); and Department of Statistics, Harvard Faculty of Arts and Sciences, 1 Oxford Street, Cambridge, Massachusetts 02138 (José R. Zubizarreta).

## Acknowledgements

The authors thank Eric Cohn (Harvard School of Public Health), Dr. Eduardo Engel (Universidad de Chile), Dr. Catterina Ferreccio (Pontificia Universidad Católica de Chile), Dr. César Hidalgo (University of Toulouse), Dr. Ronald Kessler (Harvard Medical School), Dr. Mauricio Lima (Pontificia Universidad Católica de Chile), Dr. Guillermo Marshall (Pontificia Universidad Católica de Chile), Dr. Gonzalo Mena (University of Oxford), Bijan Niknam (Harvard University), Dr. Paul Rosenbaum (University of Pennsylvania), and Dr. Zirui Song (Harvard Medical School) for helpful comments and suggestions.

## Data and material availability

The data sets used in this study are available from the Base de Datos COVID-19 repository at http://www.minciencia.gob.cl/covid19. The R code used for the analysis is available at https://scholar.harvard.edu/files/zubizarreta/files/code_v1.0.zip.

