## Supplementary material for "Effectiveness of Localized Lockdowns in the COVID-19 Pandemic": SuppMaterial_Li_etal_LocalLockdown

### Supplementary Material
### Effectiveness of Localized Lockdowns in the COVID-19 Pandemic

Yige Li,[1,2] Eduardo A. Undurraga,[3,4] José R. Zubizarreta[1,2,5,*]

[1]Department of Health Care Policy, Harvard Medical School, 180A Longwood Avenue, Boston, MA 02115, USA

[2]Department of Biostatistics, Harvard T. H. Chan School of Public Health, 677 Huntington Avenue, Boston, MA 02115, USA

[3]Escuela de Gobierno, Pontificia Universidad Católica de Chile, Vicuña Mackenna 4860, Macul, Santiago 7820436, Chile,

[4]Millennium Initiative for Collaborative Research in Bacterial Resistance (MICROB-R), Chile

[5]Department of Statistics, Harvard Faculty of Arts and Sciences, 1 Oxford Street Cambridge, MA 02138, USA

### Contents

**1 Materials and Methods** **3**

**2 Supplementary Figures** **10**

**3 Supplementary Tables** **18**

### 1 Materials and Methods

#### Estimands: direct, indirect, and total effects of lockdowns

For any given municipality $i$, we want to estimate the individual effect of lockdowns on the instantaneous reproduction number $R_{it}$ when intervening both on the duration of the lockdown $\underline{L}_{it}$ and on the proportion of the population under lockdown in the neighboring municipalities $\underline{P}_{(i)t}$. In particular, we are interested in the municipality-level direct, indirect, and total effects of lockdowns across time (*1*).

Let $\underline{l}_{it}^{a}$ and $\underline{l}_{it}^{b}$ denote two alternative lockdown interventions on municipality $i$ at time $t$, and $\underline{p}_{(i)t}^{c}$ and $\underline{p}_{(i)t}^{d}$ be two different interventions on the neighboring municipalities also at time $t$. To fix ideas, in Table S1 we provide a simple example. In the table, $\underline{l}_{it}^{a}$ denotes reopening the municipality in $t = 22$ (week 4) after two weeks of lockdown ($t = 8, ..., 21$), whereas $\underline{l}_{it}^{b}$ denotes maintaining the municipality closed during $t = 22, ..., 28$ after the same lockdown history until $t = 21$. In the table, $p_{(i)t}^{c}$ refers to reducing the population under lockdown in the neighboring municipalities to 25% from $t = 22$ after two weeks at 50%, whereas $\underline{p}_{it}^{d}$ refers to maintaining the neighboring population under lockdown at 50% after the same lockdown history.

Using this notation, for each municipality $i$, we are interested in the following causal contrasts on $R_{it}$ and $I_{it}$:

- Direct effects

$$\text{DE}_{it}^{R} = R_{it}(\underline{L}_{it} = \underline{l}_{it}^{b}, \underline{P}_{(i)t} = \underline{p}_{(i)t}^{c}) - R_{it}(\underline{L}_{it} = \underline{l}_{it}^{a}, \underline{P}_{(i)t} = \underline{p}_{(i)t}^{c}),$$

$$\text{DE}_{it}^{I} = I_{it}\left(\underline{I}_{it-1}(*), R_{it}(\underline{L}_{it} = \underline{l}_{it}^{b}, \underline{P}_{(i)t} = \underline{p}_{(i)t}^{c})\right) - I_{it}\left(\underline{I}_{it-1}(*), R_{it}(\underline{L}_{it} = \underline{l}_{it}^{a}, \underline{P}_{(i)t} = \underline{p}_{(i)t}^{c})\right);$$

- Total effects

$$\text{TE}_{it}^{R} = R_{it}(\underline{L}_{it} = \underline{l}_{it}^{b}, \underline{P}_{(i)t} = \underline{p}_{(i)t}^{d}) - R_{ijt}(\underline{L}_{it} = \underline{l}_{it}^{a}, \underline{P}_{(i)t} = \underline{p}_{(i)t}^{c}),$$

$$\text{TE}_{it}^{I} = I_{it}\left(\underline{I}_{it-1}(*), R_{it}(\underline{L}_{it} = \underline{l}_{it}^{b}, \underline{P}_{(i)t} = \underline{p}_{(i)t}^{d})\right) - I_{it}\left(\underline{I}_{it-1}(*), R_{it}(\underline{L}_{it} = \underline{l}_{it}^{a}, \underline{P}_{(i)t} = \underline{p}_{(i)t}^{c})\right);$$

and

- Indirect effects

$$\begin{aligned}
\text{IE}_{it}^{R} &= \text{TE}_{it}^{R} - \text{DE}_{it}^{R} \\
&= R_{it}(\underline{L}_{it} = \underline{l}_{it}^{b}, \underline{P}_{(i)t} = \underline{p}_{(i)t}^{d}) - R_{it}(\underline{L}_{it} = \underline{l}_{it}^{b}, \underline{P}_{(i)t} = \underline{p}_{(i)t}^{c}),
\end{aligned}$$

$$\begin{aligned}
\text{IE}_{it}^{I} &= \text{TE}_{it}^{I} - \text{DE}_{it}^{I} \\
&= I_{it}\left(\underline{I}_{it-1}(*), R_{it}(\underline{L}_{it} = \underline{l}_{it}^{b}, \underline{P}_{(i)t} = \underline{p}_{(i)t}^{d})\right) - I_{it}\left(\underline{I}_{it-1}(*), R_{it}(\underline{L}_{it} = \underline{l}_{it}^{b}, \underline{P}_{(i)t} = \underline{p}_{(i)t}^{c})\right).
\end{aligned}$$

For a group of municipalities $\mathcal{I}$, we are interested in the total effect

$$\text{TE}_{\mathcal{I}t}^{I} = \sum_{i \in \mathcal{I}} \left[ I_{it}\left(\underline{I}_{it-1}(*), R_{it}(\underline{L}_{it} = \underline{l}_{it}^{a}, \underline{P}_{(i)t} = \underline{p}_{(i)t}^{c})\right) - I_{it}\left(\underline{I}_{it-1}(*), R_{it}(\underline{L}_{it} = \underline{l}_{it}^{b}, \underline{P}_{(i)t} = \underline{p}_{(i)t}^{d})\right) \right].$$

In both the direct and total effects, one of the two potential outcomes is observed and the other one needs to be estimated.

Our estimates will be valid causal estimates under the following assumptions. First, the potential instantaneous reproduction number $R_{it}(*)$ is subject to two types of interference, direct interference and allocational interference (*2*).[1] In regards to direct interference, the instantaneous reproduction number can be approximated by the product of the time-varying transmission rate and a constant mean infectious period (*3*). The transmission rate can be considered as a function of the population under lockdown and certain characteristics of the municipality. Therefore, the effects of the intervention of one municipality on the instantaneous reproduction

[1] Under direct interference, one unit's exposure directly affects another unit's outcome, without being mediated by the first unit's outcome. Under allocational interference, one unit's outcome is affected by the units allocated to the same exposure group, either through the exposure or the outcomes of the other units in the same group (*2*).

numbers of its neighbors are not mediated by the instantaneous reproduction number in that municipality. In regards to allocational interference, we assume that the instantaneous reproduction number of one municipality cannot be affected by interventions in non-adjacent neighbors. Second, we assume that the intervention assignment is ignorable given the observed baseline covariates (*4, 5*) and the factor loadings (*6*) of the potential outcomes. Third, the decrements in the proportion of the susceptible population are negligible and the effect of the intervention is stable during our studied period. Therefore, the estimation of $R_{it}(*)$ does not differ by the calendar time.

#### Estimator: a synthetic control approach for duration and spillover effects

The basic intuition behind our approach is, for each municipality of study interest, to build a synthetic control ("clone") municipality with very similar covariate, intervention, and outcome histories until the time of the lockdown intervention by weighting appropriately in time other control municipalities that did not experience the intervention. For each municipality $i$ that is intervened at time $t$, we constructed a set of candidate control municipalities $\mathcal{C}$, as described in the next subsection. Let $\tilde{t}$ denote the last day of actual lockdown. To estimate the potential $R_{it}(*)$ during the extended lockdown, $t = \tilde{t}+1, \dots, \tilde{t}+\Delta_D$, where $\Delta_D$ represents the extension of the lockdown in days, we balanced the baseline covariates, intervention, and outcome histories with length $h$ of the control set $\mathcal{C}$ relative to those of municipality $i$. More specifically, we first estimated the potential $R_{i\tilde{t}+1}(*)$ by balancing $R_{\mathcal{C}[1,h]} := (R_{\mathcal{C}1}, \dots, R_{\mathcal{C}h})$ relative to $R_{i[\tilde{t}-h,\tilde{t}]} := (R_{i\tilde{t}-h}, \dots, R_{i\tilde{t}})$ and linearly weighting $R_{\mathcal{C}h+1}$ with the resulting balancing weights. Second, we estimated $R_{i\tilde{t}+2}(*)$ by including the estimated potential $R_{i\tilde{t}+1}(*)$ to and removing $R_{i\tilde{t}-h}$ from the history of potential $R_{i\tilde{t}+2}(*)$, and balancing $R_{\mathcal{C}[2,h+1]}$ to $R_{i[\tilde{t}-h+1,\tilde{t}+1]}(*)$. We repeated this process $\Delta_D$ times for each day in the extended lockdown period. In this way, we estimated all the potential outcomes $R_{i[\tilde{t}+1,\tilde{t}+\Delta_D]}(*) := \big(R_{i\tilde{t}+1}(*), R_{i\tilde{t}+2}(*), \dots, R_{i\tilde{t}+\Delta_D}(*)\big)$.

In this process, in addition to the history of instantaneous reproduction numbers, we also balanced the histories of lockdowns in the municipality and its neighbors and its baseline sociodemographic characteristics.

The balancing weights at time $t$ can be found by first solving a convex optimization of the form

$$\min_{\eta_{0t},\eta_{xt},\eta_{rt},\eta_{lt},\eta_{pt}} \quad \frac{1}{2}\sum_{c\in\mathcal{C}}(R_{ct_h} - (\eta_{0t} + X_c\eta_{xt} + R_{c[t_h-h,t_h-1]}\eta_{rt} + L_{c[t_h-h,t_h-1]}\eta_{lt} + P_{(c)[t_h-h,t_h-1]}\eta_{pt}))^2 \\ +\lambda_{xt}\|\eta_{xt}\|_2^2 + \lambda_{rt}\|\eta_{rt}\|_2^2 + \lambda_{lt}\|\eta_{lt}\|_2^2 + \lambda_{pt}\|\eta_{pt}\|_2^2,$$

where $t_h = t-\tilde{t}-h$; $\eta_{0t}$, $\eta_{xt}$, $\eta_{rt}$, $\eta_{lt}$ and $\eta_{pt}$ are, respectively, the intercept and vectors of regression coefficients associated with the pre-intervention baseline covariates $X_c$, time-varying reproduction numbers history $R_{c[t_h-h,t_h-1]}$ and lockdown histories $L_{c[t_h-h,t_h-1]}$ and $P_{(c)[t_h-h,t_h-1]}$; and $\lambda_{xt}$, $\lambda_{rt}$, $\lambda_{lt}$, and $\lambda_{pt}$ are ridge regression tuning parameters that control the degree of regularization and that are selected by cross-validation.

Let $\hat{\eta}_{xt}$, $\hat{\eta}_{rt}$, $\hat{\eta}_{lt}$ and $\hat{\eta}_{pt}$ be the estimated regression coefficients under $\lambda_{xt} = 0$. Let $\hat{\gamma}_{ct}$ be the implied synthetic control weights of unit $c$ at time $t$. Let $\hat{\gamma}_{\mathcal{C}t}$ define the vector of control weights $\hat{\gamma}_{ct}$. Let $R_{\mathcal{C}[t_h-h,t_h-1]}$, $L_{\mathcal{C}[t_h-h,t_h-1]}$, and $P_{\mathcal{C}[t_h-h,t_h-1]}$ denote, respectively, the matrices of $R_{c[t_h-h,t_h-1]}$, $L_{c[t_h-h,t_h-1]}$, and $P_{(c)[t_h-h,t_h-1]}$ with $|\mathcal{C}|$ rows and $h$ columns. $X_{\mathcal{C}}$ represents the matrix of baseline covariates. The augmented synthetic control weights are given by

$$\begin{aligned}
\hat{\gamma}_{ct}^{\text{cov}} \quad = \quad & \hat{\gamma}_{ct} + (\tilde{R}_{i[t-h,t-1]}(*) - \hat{\gamma}_{\mathcal{C}t}^{\top}\tilde{R}_{\mathcal{C}[t_h-h,t_h-1]})(\tilde{R}_{\mathcal{C}[t_h-h,t_h-1]}^{\top}\tilde{R}_{\mathcal{C}[t_h-h,t_h-1]} + \lambda_{rt}I_h)^{-1}\tilde{R}_{c[t_h-h,t_h-1]}^{\top} \\
& +(X_i - \hat{\gamma}_{\mathcal{C}t}^{\top}X_{\mathcal{C}})^{\top}(X_{\mathcal{C}}^{\top}X_{\mathcal{C}})^{-1}X_c^{\top} \\
& +(\tilde{l}_{i[t-h,t-1]} - \hat{\gamma}_{\mathcal{C}t}^{\top}\tilde{L}_{\mathcal{C}[t_h-h,t_h-1]})(\tilde{L}_{\mathcal{C}[t_h-h,t_h-1]}^{\top}\tilde{L}_{\mathcal{C}[t_h-h,t_h-1]} + \lambda_{lt}I_h)^{-1}\tilde{L}_{c[t_h-h,t_h-1]}^{\top} \\
& +(\tilde{p}_{(i)[t-h,t-1]} - \hat{\gamma}_{\mathcal{C}t}^{\top}\tilde{P}_{\mathcal{C}[t_h-h,t_h-1]})(\tilde{P}_{\mathcal{C}[t_h-h,t_h-1]}^{\top}\tilde{P}_{\mathcal{C}[t_h-h,t_h-1]} + \lambda_{pt}I_h)^{-1}\tilde{P}_{(c)[t_h-h,t_h-1]}^{\top},
\end{aligned}$$

where $\tilde{R}_{c[t_h-h,t_h-1]} = R_{c[t_h-h,t_h-1]} - X_c(X_{\mathcal{C}}^{\top}X_{\mathcal{C}})^{-1}X_{\mathcal{C}}^{\top}R_{\mathcal{C}[t_h-h,t_h-1]}$, $\tilde{R}_{i[t_h-h,t_h-1]}(*) = R_{i[t-h,t-1]}(*) - X_c(X_{\mathcal{C}}^{\top}X_{\mathcal{C}})^{-1}X_{\mathcal{C}}^{\top}R_{\mathcal{C}[t_h-h,t_h-1]}$, $\tilde{L}_{c[t_h-h,t_h-1]} = L_{c[t_h-h,t_h-1]} - X_c(X_{\mathcal{C}}^{\top}X_{\mathcal{C}})^{-1}X_{\mathcal{C}}^{\top}L_{\mathcal{C}[t_h-h,t_h-1]}$,

$\tilde{P}_{(c)[t_h-h,t_h-1]} = P_{(c)[t_h-h,t_h-1]} - X_c(X_{\mathcal{C}}^\top X_{\mathcal{C}})^{-1}X_{\mathcal{C}}^\top P_{\mathcal{C}[t_h-h,t_h-1]}$. $l_{i[t-h,t-1]}$ and $p_{(i)[t-h,t-1]}$ are intervention histories, $\tilde{l}_{i[t-h,t-1]}$, $\tilde{p}_{(i)[t-h,t-1]}$ are projected into $X_{\mathcal{C}}$ similarly. The weights $\hat{\gamma}_{ct}^{\text{cov}}$ exactly balance baseline covariates and approximately balance lagged interventions and outcomes. Point estimates of $R_{it}(*)$ are produced by linearly weighting the observed outcomes.

The weights are not constrained to take non-negative values; hence the estimated instantaneous reproduction number can take values that are not sample bounded. We note, however, the regularization terms in the optimization problem prevent this from happening and the weights can be checked empirically. (In our study, we checked this, and the estimated reproduction number took valid numbers.)

Following (*7*), we computed $I_{it}(*)$ from the estimated $R_{it}(*)$ using the unbiased estimator $I_{it}(*) = R_{it}(*)\sum_{s=1}^{t} I_{it-s}(*)w_s$. Therefore, $\text{Var}(I_{it}(*)|\{I_{it-s}(*)\}_{s=1}^{t}) = \left(\sum_{s=1}^{t} I_{it-s}(*)w_s\right)^2 \text{Var}(R_{it}(*))$. See (*6*) for details of the variance of the resulting estimator.

#### Further details and code

In our estimations, we set $h = 7$ because the partial autocorrelations of the estimated $R_{it}$ for lags over 7 days is negligible in our data set. We also assumed that $R_{it}$ is not affected by the histories of lockdown interventions and proportion of neighboring population under lockdown beyond 7 days.

For each intervention municipality $i$, we built a set $\mathcal{C}$ of candidate control municipality-periods as follows. We considered municipalities that had at least 10 reported cases and were under lockdown for at least 14 days during the study period. For each of these candidate control municipalities $k \neq i$, we built a subset $\mathcal{C}_k$ of municipality-periods of 14 contiguous days from the start of its lockdown intervention. Therefore, a candidate control municipality $k$ that was under lockdown for 14 days contributes one municipality-period to $\mathcal{C}_k$, a control municipality $k'$ that was under lockdown for 15 days contributes 2 municipality-periods to $\mathcal{C}'_k$, and so on. In general, a candidate control municipality $k$ that was under lockdown for $D_k$ days contributes $D_k - 14 + 1$ control municipality-periods to $\mathcal{C}_k$. $\mathcal{C}$ is the union of all the sets $\mathcal{C}_k$.

After estimating all the potential $R_{i[\tilde{t}+1,\tilde{t}+7]}(*)$ in the first week of extended lockdown, we used those predicted values to repeat the balancing process in the second week, and similarly in the third week. We refrained from providing predictions for more weeks as the prediction errors increase and the estimates become more unstable.

Assuming that the error terms of $R_{ct}$ of all the units in the control set are independent across $c$ and $t$ and follow a sub-Gaussian distribution, the estimated conditional variance of each estimation from $\tilde{t}+1$ to $\tilde{t}+\Delta_D$ given its history was obtained using the R package of (*6*). The variance of estimation at each time was calculated by the law of total variance.

To estimate the variance of the average $R_{it}(*)$ from $\tilde{t}+1$ to $\tilde{t}+\Delta_D$, we first estimated the autocorrelation of the observed series in $i$ and then estimated the variance of the sum of those correlated random variables.

We conducted all our analyses using `R`. The code can be found through this link: `https://scholar.harvard.edu/files/zubizarreta/files/code_v1.0.zip`.

#### Baseline covariates and lagged variables

Following (*3*), the instantaneous reproduction number is proportional to the time-varying transmission rate, which is a function of certain characteristics of the municipalities. We considered both baseline (time-invariant) covariates and lagged (time-varying) variables. Both of these variables may confound the effect of the lockdown interventions, so we adjusted for them. A summary of the baseline, time-invariant covariates is in Table S2. The lagged variables include: $h$-day history of instantaneous reproduction number, $h$-day history of proportion of neighboring population under lockdown, and $h$-day history of lockdown interventions.

#### Data adjustments

We adjusted the COVID-19 case incidence series to correct the lag in reporting and some incomplete municipality-level data. First, we imputed the incomplete data by interpolating between the closest dates with complete data. In the data, the number of cumulative cases was reported typically every 2-4 days. We interpolated between the closest dates to acquired daily new cases respectively by reporting date and symptoms onset date. Second, we estimated the lags in reporting using the PELT algorithm (*8*). In the data, the number of cases showing symptoms was reported weekly. We smoothed these values by computing daily moving averages with a window of 7 days. For the reported cases on each date, we first estimated their reporting lags by finding their symptoms onset date based on the fact that almost all the cases were traced for their symptom onset date. After that, we assumed that the reporting lags followed a Poisson distribution with a time-varying expected rates of occurrences. To avoid overfitting, the number of change points in the expected rates and the corresponding values of those rates were found by the PELT algorithm (*8*) with a penalty for regularization. Now, for the reported cases on each date, we had an estimated reporting lag. Third, we estimated the adjusted incidence series and instantaneous reproduction number series employing the implementation by Zhao et al. (*9*) of Cori et al. (*7*). In each of the 1000 replication, the reporting lags were drawn from the Poisson distribution with time-varying rates. In summary, these adjustments considered that the lag between symptom onset date and report date could vary across municipalities and over time.

#### Data and code availability

The data sets used and/or analyzed in this study are available from Base de Datos COVID-19 repository at `http://www.minciencia.gob.cl/covid19`. The R code used for analysis is available at `https://scholar.harvard.edu/files/zubizarreta/files/code_v1.0.zip`.

### 2 Supplementary Figures

#### Duration of the lockdowns and proportion of neighbors under lockdown

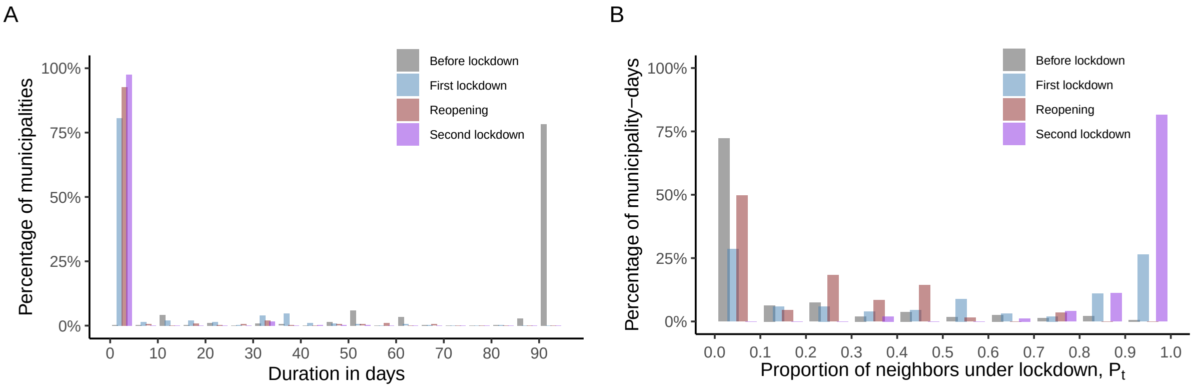


**Figure S1: Background for Figure 1**. **A:** Histogram of the durations of different lockdown statuses in all municipalities during the study period (before lockdown, during the first lockdown, after the reopening, during the second lockdown). From March 15 to June 15, all of the lockdown interventions occur within 21.7% of the municipalities. The rest of the municipalities did not experience lockdowns by the end of the study period. The median duration of the first lockdown is 32 days. **B:** Histogram of the proportion of neighbors under lockdown, $P_t$, in municipality-days from March 15 to June 15. During the second lockdown period, 81.5% of the municipality-days showed more than 90.0% of its neighboring population under lockdown.

### Imbalances of lagged variables

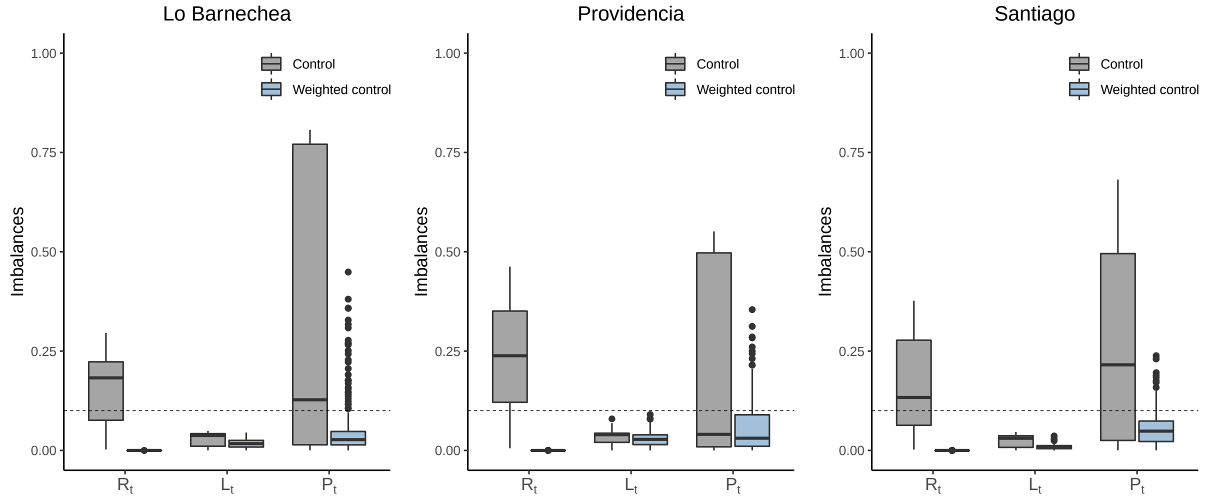


**Figure S2: Imbalances of lagged variables of municipalities Lo Barnechea, Providencia, and Santiago in analysis in Figure 2**. Imbalances of all the lagged variables, including the instantaneous reproduction number $R_t$, the lockdown indicator $L_t$, and the proportion of neighboring population under lockdown $P_t$, in the control units before and after weighting. The dashed line at 0.1 denotes the commonly accepted balance threshold. After weighting, the imbalances of $R_t$ is near 0, the imbalance of $L_t$ is reduced substantially, and the third quartile of the imbalance of $P_t$ is below 0.1.

### Results for other municipalities in Greater Santiago

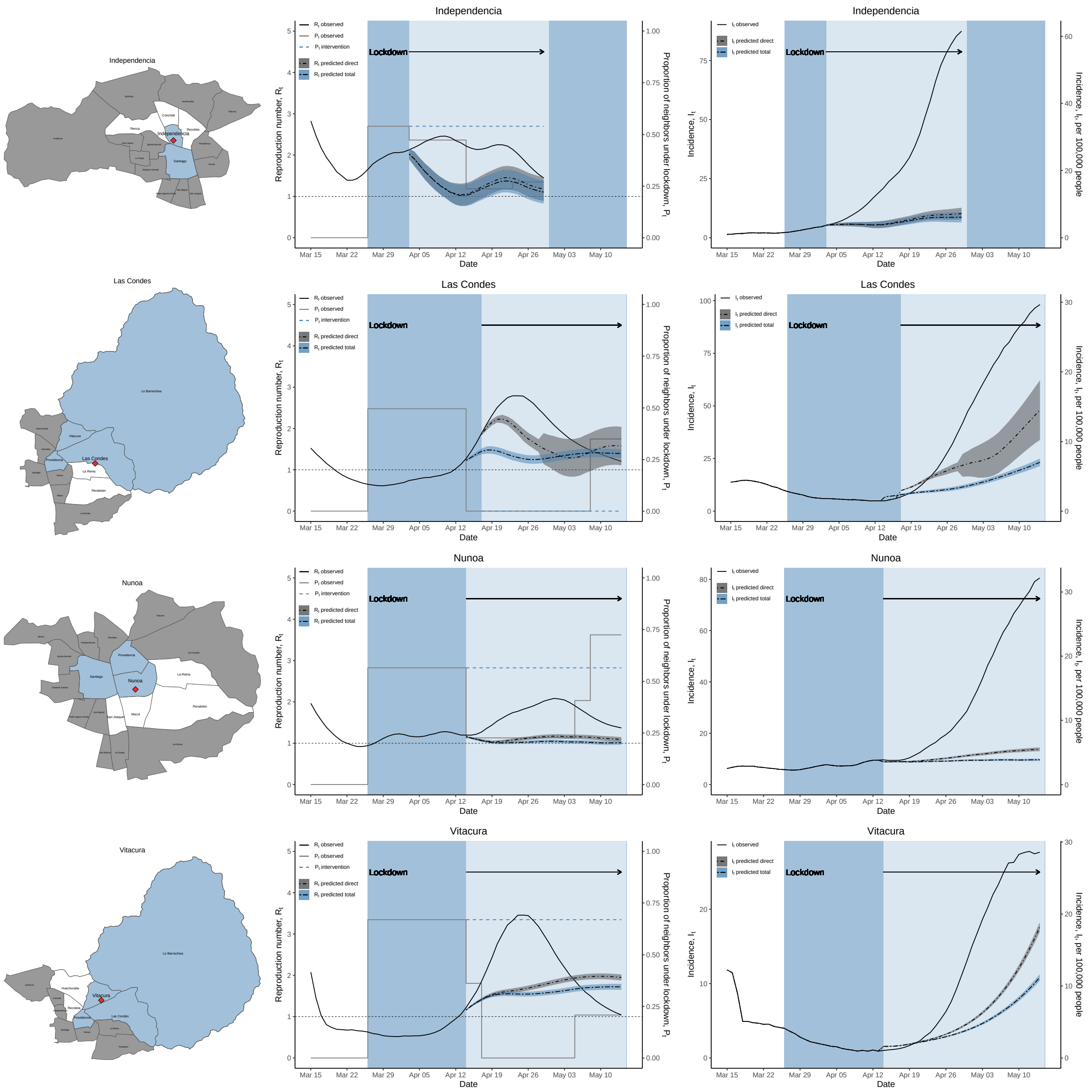


**Figure S3: Results for extended lockdown in municipalities Independencia, Las Condes, Ñuñoa, and Vitacura with the same analysis in Figure 2**. The shaded error bands are the 95% confidence intervals based on augmented synthetic control method. The exact numbers are in Table S7 and S8. The analysis for the total effect on Las Condes is slightly different from the otehrs. The neighbors of Las Condes reopened simultaneously three days before its own reopening. To study the total effect, we intervene on its neighbors Providencia, Vitacura, and Lo Barnechea to maintain their lockdown status rather than reopen.

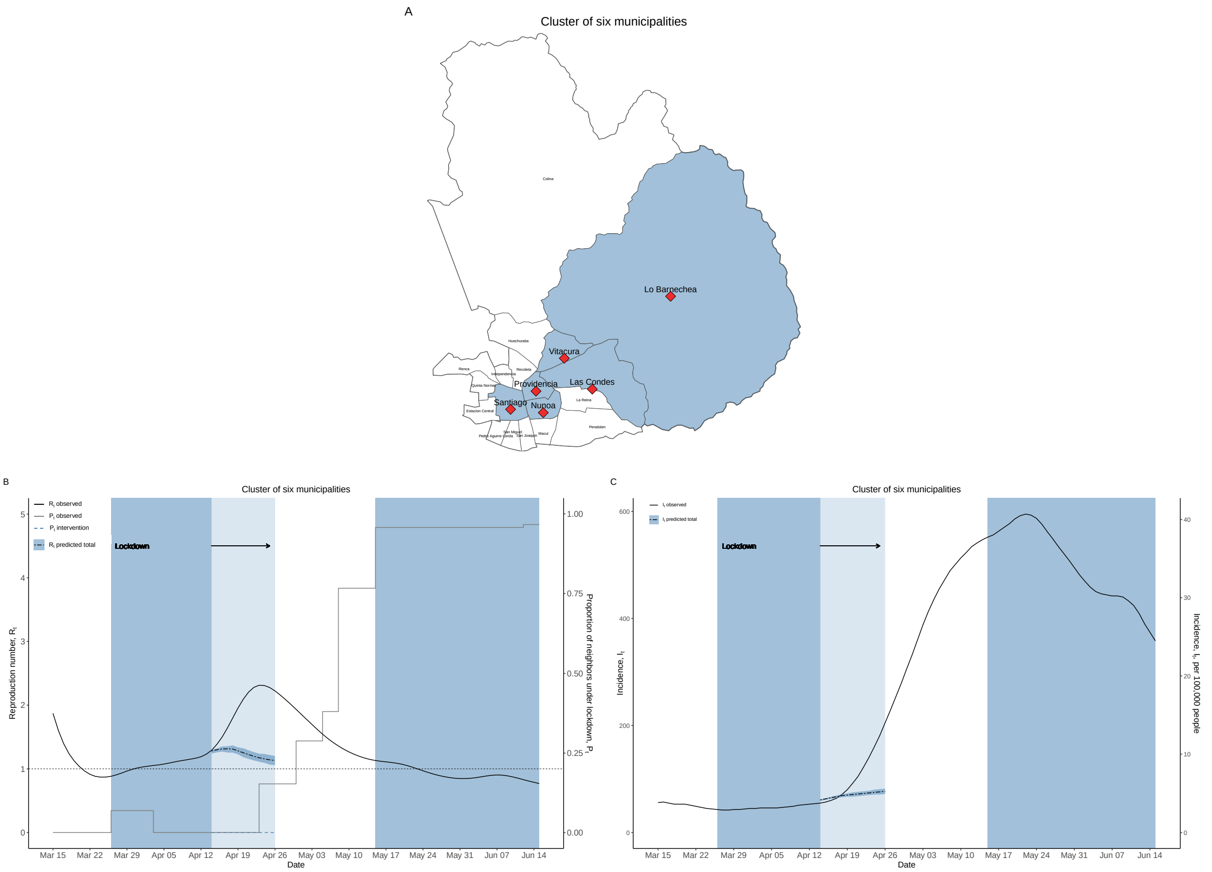


**Figure S4: Results for extended lockdown in a cluster of six municipalities in Greater Santiago**. **A**. No neighbors of the cluster of six municipalities are under lockdown on April 13. **B**. The estimates of the potential $R_t(*)$ if the lockdown is extended from March 26 to April 26, when their neighbors remain open. **C**. The estimates of potential $I_t(*)$. The second lockdown period from May 15 to June 15 is included as a comparison to the first lockdown period from March 26 to April 26.

### Results for other municipalities in the rest of Chile

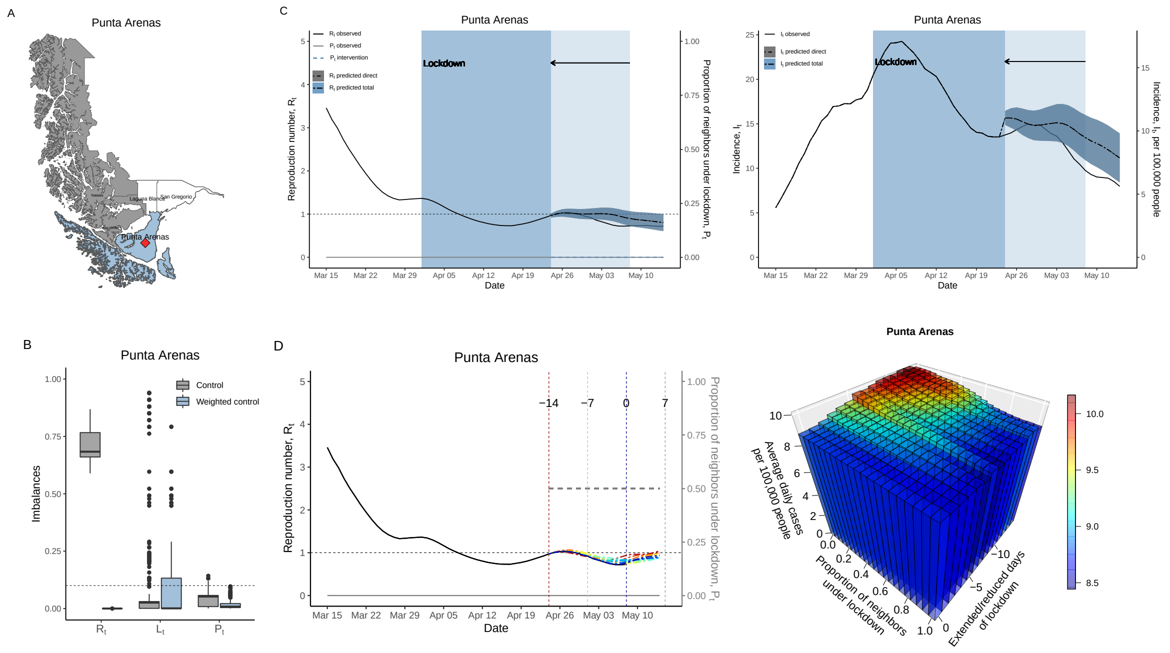


**Figure S5: Results for Punta Arenas**. **A**. Punta Arenas is a relatively isolated municipality. **B**. Imbalances in the lagged variables before and after weighting.[2] **C**. The initial weeks of lockdown in Punta Arenas were effective in terms of reducing the instantaneous reproduction number and the incidence. After reopening, the reproduction number stabilized at a low level. The exact numbers are in Table S10. **D**. The impact of neighboring lockdowns on Punta Arenas is relatively small. The exact numbers are in Table S14.

[2]To estimate the impact of reducing lockdown duration, the control set is composed of candidate municipalities which were under lockdown for at least 7 days and had reopened for at least 14 days.

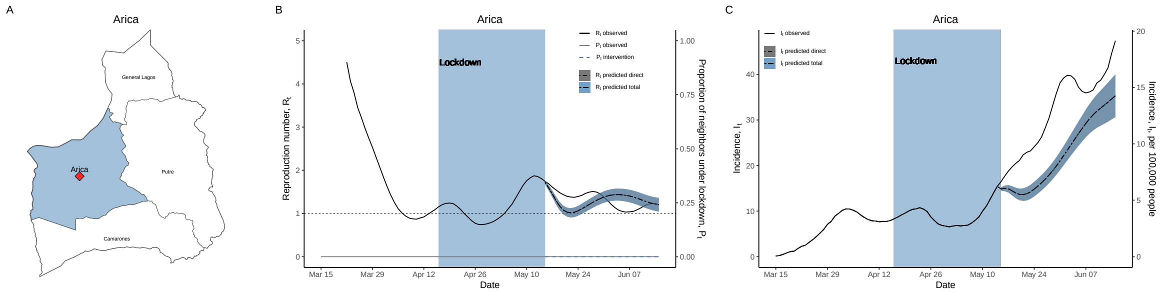


**Figure S6: Results for Arica as a comparison to Punta Arenas**. **A**. Arica is another relatively isolated municipality. **B**. During lockdown, the average reproduction number is above one. After reopening, both the actual reproduction numbers and the predicted ones are also above one. **C**. Both the predicted and actual series exhibit a steady increase in the incidence.

### Estimated direct and total effects over time

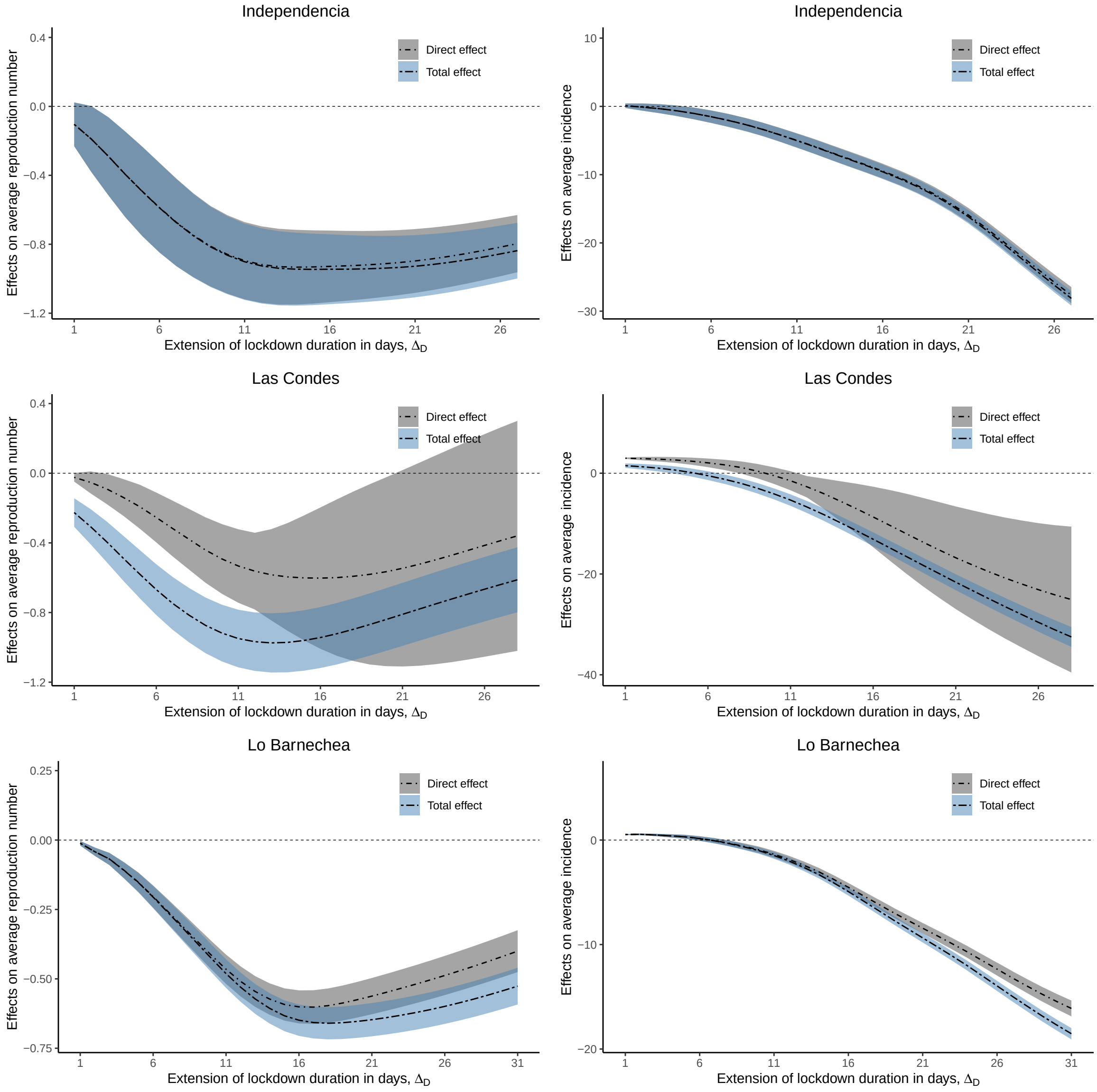


**Figure S7:** Direct and total effect of extended lockdowns on average reproduction number $\text{avg}(R_{[\tilde{t}+1,\tilde{t}+\Delta_D]}(*))$ and average incidence $\text{avg}(I_{[\tilde{t}+1,\tilde{t}+\Delta_D]}(*))$ in Independencia, Las Condes, and Lo Barnechea. The shaded error bands are the 95% confidence intervals. The exact numbers are in Table S9.

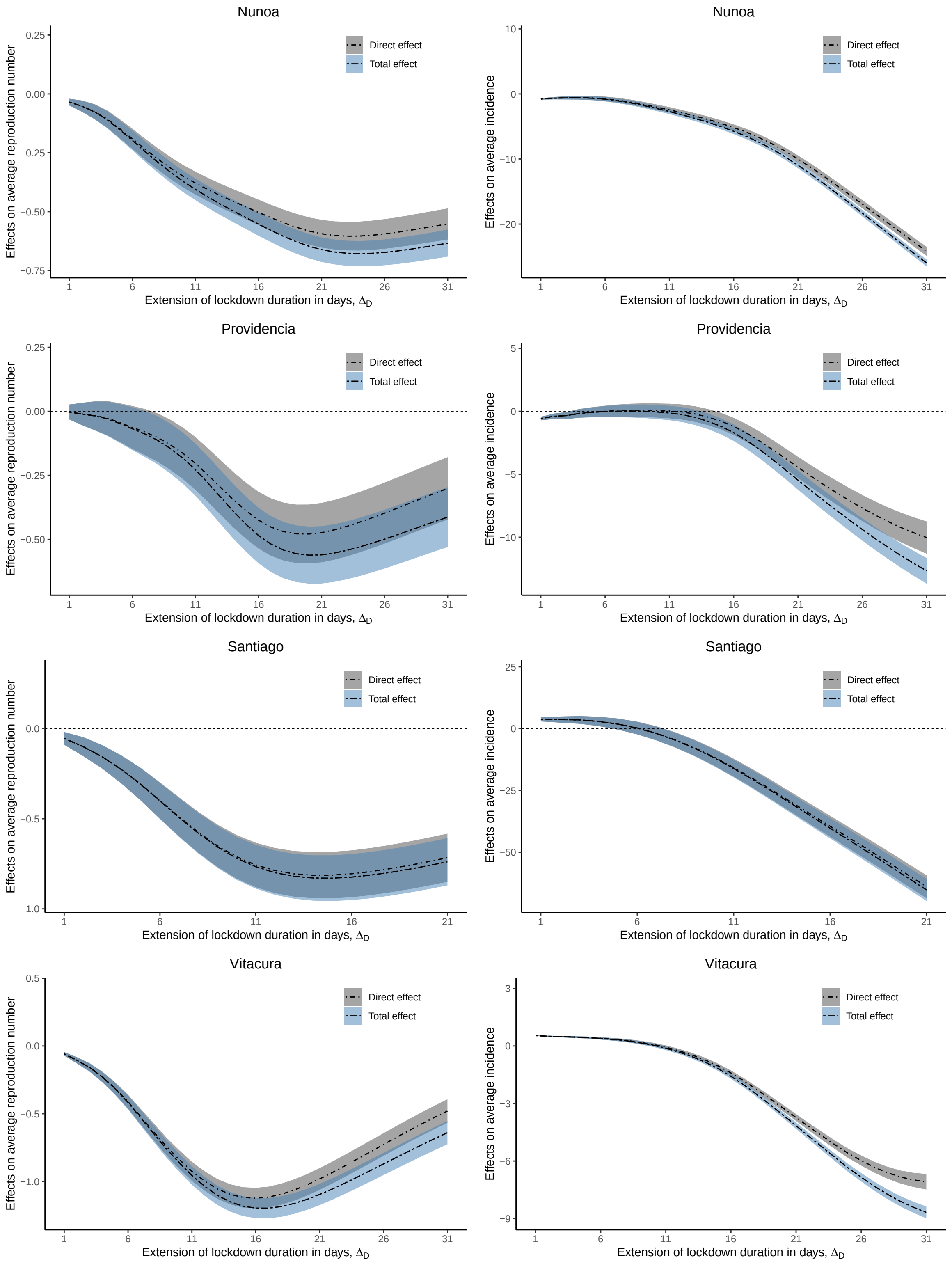


**Figure S8:** Direct and total effects of extended lockdowns in Ñuñoa, Providencia, Santiago, and Vitacura.

### 3 Supplementary Tables

#### An example

**Table S1:** Example alternative lockdown interventions at the level of the municipality and its neighbors over a time horizon of 4 weeks.

| Intervention | Time | | | |
|---|---|---|---|---|
| | $t = 1, ..., 7$ | $t = 8, ..., 14$ | $t = 15, ..., 21$ | $t = 22, ..., 28$ |
| On municipality $i$ | | | | |
| $\underline{l}_{it}^{a}$ | 0 | 1 | 1 | 0 |
| $\underline{l}_{it}^{b}$ | 0 | 1 | 1 | 1 |
| On the neighbors of municipality $i$ | | | | |
| $\underline{p}_{(i)t}^{c}$ | 0 | 0.5 | 0.5 | 0.25 |
| $\underline{p}_{(i)t}^{d}$ | 0 | 0.5 | 0.5 | 0.5 |

#### Baseline covariates

**Table S2:** Summary of baseline covariates in 324 municipalities with no missing values.

| Variable | Mean (SD) | IQR (25%, 75%) | Description |
|---|---|---|---|
| Rural | 0.37 (0.28) | 0.09, 0.59 | Proportion of the population in rural areas. |
| Female | 0.52 (0.03) | 0.50, 0.53 | Proportion of female population. |
| Over 65 | 0.13 (0.03) | 0.11, 0.15 | Proportion of the population over 65. |
| Poverty | 0.13 (0.08) | 0.07, 0.17 | Proportion of the population living with a monthly income per capita below the poverty line (about 3.9 USD per person per day in 2017). |
| Overcrowding | 0.09 (0.05) | 0.06, 0.12 | Proportion of households with an overcrowding condition (people/rooms $\geq 2.5$). |
| Poor sanitation | 0.14 (0.15) | 0.04, 0.20 | Proportion of the population with inadequate sanitation (House Sanitation Index). Inadequate sanitation defined as a household with no access to drinkable water, or without a toilet or latrine connected to the sewer or septic tank. |
| Income | 285691.44 (152089.93) | 212992.25, 305508.25 | Average income per capita in 2017 CLP (Chilean pesos; 1 USD = 700 CLP). |
| Area | 1966.99 (4758.30) | 243.75, 1455.95 | Area in $km^2$. Area is divided into small/medium/large area in the balancing procedure. |

#### Covariate balance

**Table S3: Balance in baseline covariates and lagged variables**. Balance in the municipality of Lo Barnechea and its control set at the start of the intervention $\tilde{t}+1$, before and after weighting. See also Figure S2 for a summary distribution of imbalances in all the lagged variables.

| Baseline covariates | Lo Barnechea | Control Set | Synthetic Lo Barnechea |
|---|---|---|---|
| Rural | 0.00 | 0.03 | 0.00 |
| Female | 0.56 | 0.52 | 0.56 |
| Over 65 | 0.08 | 0.11 | 0.08 |
| Poverty | 0.03 | 0.07 | 0.03 |
| Overcrowding | 0.12 | 0.12 | 0.12 |
| Poor sanitation | 0.05 | 0.04 | 0.05 |
| Income | 714251.33 | 378907.93 | 714251.33 |
| Area (small) | 0.00 | 0.35 | 0.00 |
| Area (medium) | 1.00 | 0.36 | 1.00 |
| Area (large) | 0.00 | 0.29 | 0.00 |
| Lagged variables | Lo Barnechea | Control Set | Synthetic Lo Barnechea |
| $R_{t-7}$ | 1.43 | 1.14 | 1.43 |
| $R_{t-6}$ | 1.41 | 1.13 | 1.41 |
| $R_{t-5}$ | 1.38 | 1.12 | 1.38 |
| $R_{t-4}$ | 1.33 | 1.10 | 1.33 |
| $R_{t-3}$ | 1.29 | 1.10 | 1.29 |
| $R_{t-2}$ | 1.27 | 1.09 | 1.27 |
| $R_{t-1}$ | 1.28 | 1.08 | 1.28 |
| $L_{t-7}$ | 1.00 | 0.95 | 1.03 |
| $L_{t-6}$ | 1.00 | 0.95 | 1.03 |
| $L_{t-5}$ | 1.00 | 0.95 | 1.01 |
| $L_{t-4}$ | 1.00 | 0.95 | 1.00 |
| $L_{t-3}$ | 1.00 | 0.96 | 0.99 |
| $L_{t-2}$ | 1.00 | 0.96 | 0.97 |
| $L_{t-1}$ | 1.00 | 0.96 | 0.96 |
| $P_{t-7}$ | 0.53 | 0.73 | 0.52 |
| $P_{t-6}$ | 0.53 | 0.74 | 0.53 |
| $P_{t-5}$ | 0.53 | 0.75 | 0.53 |
| $P_{t-4}$ | 0.53 | 0.75 | 0.54 |
| $P_{t-3}$ | 0.53 | 0.76 | 0.54 |
| $P_{t-2}$ | 0.53 | 0.77 | 0.52 |
| $P_{t-1}$ | 0.53 | 0.78 | 0.54 |

**Table S4: Balance in baseline covariates and lagged variables**. Balance in the municipality of Providencia and its control set at the start of the intervention $\tilde{t}+1$, before and after weighting. See also Figure S2 for a summary distribution of imbalances in all the lagged variables.

| Baseline covariates | Providencia | Control Set | Synthetic Providencia |
|---|---|---|---|
| Rural | 0.00 | 0.03 | 0.00 |
| Female | 0.53 | 0.52 | 0.53 |
| Over 65 | 0.16 | 0.11 | 0.16 |
| Poverty | 0.00 | 0.07 | 0.00 |
| Overcrowding | 0.01 | 0.12 | 0.01 |
| Poor sanitation | 0.01 | 0.04 | 0.01 |
| Income | 1360119.37 | 375505.64 | 1360119.37 |
| Area (small) | 1.00 | 0.35 | 1.00 |
| Area (medium) | 0.00 | 0.36 | 0.00 |
| Area (large) | 0.00 | 0.29 | 0.00 |
| Lagged variables | Providencia | Control Set | Synthetic Providencia |
| $R_{t-7}$ | 1.41 | 1.14 | 1.41 |
| $R_{t-6}$ | 1.42 | 1.13 | 1.42 |
| $R_{t-5}$ | 1.41 | 1.11 | 1.41 |
| $R_{t-4}$ | 1.36 | 1.10 | 1.36 |
| $R_{t-3}$ | 1.31 | 1.09 | 1.31 |
| $R_{t-2}$ | 1.24 | 1.09 | 1.24 |
| $R_{t-1}$ | 1.17 | 1.08 | 1.17 |
| $L_{t-7}$ | 1.00 | 0.95 | 1.03 |
| $L_{t-6}$ | 1.00 | 0.95 | 1.02 |
| $L_{t-5}$ | 1.00 | 0.95 | 1.01 |
| $L_{t-4}$ | 1.00 | 0.95 | 1.00 |
| $L_{t-3}$ | 1.00 | 0.96 | 0.99 |
| $L_{t-2}$ | 1.00 | 0.96 | 0.98 |
| $L_{t-1}$ | 1.00 | 0.96 | 0.96 |
| $P_{t-7}$ | 0.80 | 0.73 | 0.82 |
| $P_{t-6}$ | 0.80 | 0.74 | 0.81 |
| $P_{t-5}$ | 0.80 | 0.74 | 0.81 |
| $P_{t-4}$ | 0.80 | 0.75 | 0.81 |
| $P_{t-3}$ | 0.80 | 0.76 | 0.80 |
| $P_{t-2}$ | 0.80 | 0.77 | 0.79 |
| $P_{t-1}$ | 0.80 | 0.78 | 0.78 |

**Table S5: Balance in baseline covariates and lagged variables**. Balance in the municipality of Santiago and its control set at the start of the intervention $\tilde{t} + 1$, before and after weighting. See also Figure S2 for a summary distribution of imbalances in all the lagged variables.

| Baseline covariates | Santiago | Control Set | Synthetic Santiago |
|---|---|---|---|
| Rural | 0.00 | 0.03 | 0.00 |
| Female | 0.50 | 0.52 | 0.50 |
| Over 65 | 0.07 | 0.11 | 0.07 |
| Poverty | 0.04 | 0.07 | 0.04 |
| Overcrowding | 0.19 | 0.12 | 0.19 |
| Poor sanitation | 0.02 | 0.04 | 0.02 |
| Income | 593633.86 | 375521.94 | 593633.86 |
| Area (small) | 1.00 | 0.36 | 1.00 |
| Area (medium) | 0.00 | 0.35 | 0.00 |
| Area (large) | 0.00 | 0.30 | 0.00 |
| Lagged variables | Santiago | Control Set | Synthetic Santiago |
| $R_{t-7}$ | 1.14 | 1.13 | 1.14 |
| $R_{t-6}$ | 1.14 | 1.12 | 1.14 |
| $R_{t-5}$ | 1.14 | 1.10 | 1.14 |
| $R_{t-4}$ | 1.17 | 1.09 | 1.17 |
| $R_{t-3}$ | 1.19 | 1.08 | 1.19 |
| $R_{t-2}$ | 1.24 | 1.07 | 1.24 |
| $R_{t-1}$ | 1.31 | 1.07 | 1.31 |
| $L_{t-7}$ | 1.00 | 0.95 | 1.01 |
| $L_{t-6}$ | 1.00 | 0.95 | 1.01 |
| $L_{t-5}$ | 1.00 | 0.96 | 1.00 |
| $L_{t-4}$ | 1.00 | 0.96 | 1.00 |
| $L_{t-3}$ | 1.00 | 0.96 | 1.00 |
| $L_{t-2}$ | 1.00 | 0.96 | 0.99 |
| $L_{t-1}$ | 1.00 | 0.96 | 0.99 |
| $P_{t-7}$ | 0.36 | 0.74 | 0.35 |
| $P_{t-6}$ | 0.36 | 0.75 | 0.33 |
| $P_{t-5}$ | 0.36 | 0.76 | 0.34 |
| $P_{t-4}$ | 0.36 | 0.76 | 0.36 |
| $P_{t-3}$ | 0.36 | 0.77 | 0.38 |
| $P_{t-2}$ | 0.36 | 0.78 | 0.36 |
| $P_{t-1}$ | 0.36 | 0.79 | 0.39 |

**Table S6: Balance in baseline covariates and lagged variables**. Balance in the municipality of Punta Arenas and its control set at the start of intervention, before and after weighting. See also Figure S5B for a summary distribution of imbalances in all the lagged variables.

| Baseline covariates | Punta Arenas | Control Set | Synthetic Punta Arenas |
|---|---|---|---|
| Rural | 0.06 | 0.14 | 0.06 |
| Female | 0.52 | 0.53 | 0.52 |
| Over 65 | 0.12 | 0.12 | 0.12 |
| Poverty | 0.02 | 0.11 | 0.02 |
| Overcrowding | 0.07 | 0.09 | 0.07 |
| Poor sanitation | 0.06 | 0.09 | 0.06 |
| Income | 492972.66 | 474566.28 | 492972.66 |
| Area (small) | 0.00 | 0.36 | 0.00 |
| Area (medium) | 0.00 | 0.38 | 0.00 |
| Area (large) | 1.00 | 0.26 | 1.00 |
| Lagged variables | Punta Arenas | Control Set | Synthetic Punta Arenas |
| $R_{t-7}$ | 0.73 | 1.54 | 0.73 |
| $R_{t-6}$ | 0.75 | 1.56 | 0.75 |
| $R_{t-5}$ | 0.77 | 1.58 | 0.77 |
| $R_{t-4}$ | 0.80 | 1.60 | 0.80 |
| $R_{t-3}$ | 0.84 | 1.61 | 0.84 |
| $R_{t-2}$ | 0.88 | 1.62 | 0.88 |
| $R_{t-1}$ | 0.92 | 1.63 | 0.92 |
| $L_{t-7}$ | 1.00 | 0.24 | 1.28 |
| $L_{t-6}$ | 1.00 | 0.21 | 1.27 |
| $L_{t-5}$ | 1.00 | 0.18 | 1.21 |
| $L_{t-4}$ | 1.00 | 0.15 | 1.10 |
| $L_{t-3}$ | 1.00 | 0.12 | 0.94 |
| $L_{t-2}$ | 1.00 | 0.09 | 0.73 |
| $L_{t-1}$ | 1.00 | 0.06 | 0.48 |
| $P_{t-7}$ | 0.00 | 0.14 | 0.06 |
| $P_{t-6}$ | 0.00 | 0.13 | 0.04 |
| $P_{t-5}$ | 0.00 | 0.12 | 0.02 |
| $P_{t-4}$ | 0.00 | 0.11 | 0.00 |
| $P_{t-3}$ | 0.00 | 0.10 | -0.02 |
| $P_{t-2}$ | 0.00 | 0.09 | -0.04 |
| $P_{t-1}$ | 0.00 | 0.08 | -0.06 |

#### Estimates

**Table S7: Numeric results in Figure 2D and Figure S3**. Observed and estimated instantaneous reproduction numbers when the duration of the lockdown in a given municipality is extended from date $\tilde{t}$ to $\tilde{t} + \Delta_D$, and when in addition to extending the duration of the lockdown, the proportion of the population under lockdown in the neighboring municipalities at date $\tilde{t}$ is maintained constant until $\tilde{t} + \Delta_D$.

| | $\Delta_D$ | $P_{[\tilde{t}+1,\tilde{t}+\Delta_D]}$ | $R_{\tilde{t}+7}(*)$ | $R_{\tilde{t}+14}(*)$ | $R_{\tilde{t}+21}(*)$ |
|---|---|---|---|---|---|
| Independencia | 0 | $p^{\text{obs}}$ | 2.45 | 2.13 | 2.14 |
| Independencia | 21 | $p^{\text{obs}}$ | 1.27 [1.05, 1.48] | 1.16 [0.89, 1.43] | 1.44 [1.15, 1.72] |
| Independencia | 21 | 54.0% | 1.26 [1.05, 1.47] | 1.12 [0.86, 1.39] | 1.35 [1.08, 1.63] |
| Las Condes | 0 | $p^{\text{obs}}$ | 2.79 | 2.21 | 1.53 |
| Las Condes | 21 | $p^{\text{obs}}$ | 2.09 [1.97, 2.22] | 1.45 [1.07, 1.84] | 1.38 [0.92, 1.84] |
| Las Condes | 21 | 49.5%[3] | 1.46 [1.36, 1.55] | 1.25 [1.14, 1.35] | 1.36 [1.26, 1.47] |
| Lo Barnechea | 0 | $p^{\text{obs}}$ | 1.83 | 2.17 | 1.62 |
| Lo Barnechea | 21 | $p^{\text{obs}}$ | 1.26 [1.23, 1.29] | 1.24 [1.20, 1.28] | 1.32 [1.27, 1.36] |
| Lo Barnechea | 21 | 53.0% | 1.24 [1.21, 1.27] | 1.12 [1.09, 1.16] | 1.10 [1.06, 1.14] |
| Ñuñoa | 0 | $p^{\text{obs}}$ | 1.54 | 1.90 | 1.98 |
| Ñuñoa | 21 | $p^{\text{obs}}$ | 1.04 [1.00, 1.07] | 1.14 [1.09, 1.18] | 1.16 [1.11, 1.21] |
| Ñuñoa | 21 | 56.5% | 1.01 [0.98, 1.04] | 1.03 [0.99, 1.07] | 1.03 [0.99, 1.08] |
| Providencia | 0 | $p^{\text{obs}}$ | 1.39 | 2.47 | 1.85 |
| Providencia | 21 | $p^{\text{obs}}$ | 1.20 [1.14, 1.26] | 1.51 [1.43, 1.58] | 1.48 [1.40, 1.56] |
| Providencia | 21 | 80.3% | 1.18 [1.11, 1.24] | 1.37 [1.30, 1.43] | 1.31 [1.24, 1.38] |
| Santiago | 0 | $p^{\text{obs}}$ | 2.34 | 2.03 | 1.46 |
| Santiago | 21 | $p^{\text{obs}}$ | 1.30 [1.23, 1.38] | 1.12 [1.03, 1.20] | 1.18 [1.09, 1.27] |
| Santiago | 21 | 35.8% | 1.29 [1.22, 1.37] | 1.08 [0.99, 1.16] | 1.16 [1.07, 1.24] |
| Vitacura | 0 | $p^{\text{obs}}$ | 2.76 | 3.33 | 2.01 |
| Vitacura | 21 | $p^{\text{obs}}$ | 1.57 [1.51, 1.62] | 1.72 [1.66, 1.78] | 1.91 [1.85, 1.98] |
| Vitacura | 21 | 66.9% | 1.54 [1.49, 1.59] | 1.55 [1.49, 1.61] | 1.65 [1.58, 1.71] |

[3] To be consistent with the rest of the municipalities in the group, $p$ is the value of $P_t$ on the last day before reopening in Lo Barnechea, Vitacura, Providencia, Santiago, and Ñuñoa.

**Table S8: Numeric results in Figure 2E and Figure S3**. Observed and estimated incidences when the duration of the lockdown in a given municipality is extended from date $\tilde{t}$ to $\tilde{t}+\Delta_D$, and when in addition to extending the duration of the lockdown, the proportion of the population under lockdown in the neighboring municipalities at date $\tilde{t}$ is maintained constant until $\tilde{t}+\Delta_D$.

| | $\Delta_D$ | $P_{[\tilde{t}+1,\tilde{t}+\Delta_D]}$ | $I_{\tilde{t}+7}(*)$ | $I_{\tilde{t}+14}(*)$ | $I_{\tilde{t}+21}(*)$ |
|---|---|---|---|---|---|
| Independencia | 0 | $p^{\text{obs}}$ | 10.93 | 25.70 | 58.10 |
| Independencia | 21 | $p^{\text{obs}}$ | 5.66 [4.71, 6.62] | 6.39 [4.88, 7.89] | 9.39 [7.53, 11.25] |
| Independencia | 21 | 54.0% | 5.63 [4.68, 6.58] | 6.13 [4.68, 7.57] | 8.46 [6.74, 10.19] |
| Las Condes | 0 | $p^{\text{obs}}$ | 17.08 | 47.49 | 77.65 |
| Las Condes | 21 | $p^{\text{obs}}$ | 16.35 [15.37, 17.32] | 22.53 [16.52, 28.55] | 30.10 [20.08, 40.12] |
| Las Condes | 21 | 49.5% | 9.49 [8.78, 10.20] | 12.01 [11.04, 12.98] | 16.87 [15.60, 18.14] |
| Lo Barnechea | 0 | $p^{\text{obs}}$ | 8.91 | 20.49 | 35.86 |
| Lo Barnechea | 21 | $p^{\text{obs}}$ | 7.62 [7.43, 7.81] | 9.41 [9.1, 9.71] | 12.40 [11.95, 12.84] |
| Lo Barnechea | 21 | 53.0% | 7.54 [7.36, 7.72] | 8.35 [8.08, 8.62] | 9.12 [8.78, 9.45] |
| Ñuñoa | 0 | $p^{\text{obs}}$ | 11.43 | 21.13 | 46.42 |
| Ñuñoa | 21 | $p^{\text{obs}}$ | 9.10 [8.81, 9.39] | 10.47 [10.06, 10.88] | 12.10 [11.57, 12.64] |
| Ñuñoa | 21 | 56.5% | 8.84 [8.57, 9.11] | 9.21 [8.87, 9.55] | 9.52 [9.12, 9.92] |
| Providencia | 0 | $p^{\text{obs}}$ | 6.40 | 13.91 | 33.95 |
| Providencia | 21 | $p^{\text{obs}}$ | 6.75 [6.39, 7.11] | 10.32 [9.83, 10.82] | 15.05 [14.27, 15.82] |
| Providencia | 21 | 80.3% | 6.62 [6.26, 6.98] | 9.10 [8.64, 9.57] | 11.71 [11.07, 12.36] |
| Santiago | 0 | $p^{\text{obs}}$ | 52.37 | 133.64 | 211.34 |
| Santiago | 21 | $p^{\text{obs}}$ | 37.50 [35.27, 39.73] | 40.65 [37.51, 43.79] | 48.47 [44.84, 52.11] |
| Santiago | 21 | 35.8% | 37.17 [34.97, 39.36] | 38.83 [35.79, 41.87] | 45.38 [41.97, 48.80] |
| Vitacura | 0 | $p^{\text{obs}}$ | 1.98 | 7.56 | 20.20 |
| Vitacura | 21 | $p^{\text{obs}}$ | 2.02 [1.95, 2.09] | 3.47 [3.35, 3.60] | 6.70 [6.47, 6.93] |
| Vitacura | 21 | 66.9% | 1.98 [1.91, 2.05] | 3.04 [2.93, 3.16] | 5.01 [4.81, 5.20] |

**Table S9: Numeric results in Figure S7, S8**. Observed and estimated average instantaneous reproduction number and cumulative incidences when the duration of the lockdown in a given municipality is extended from date $\tilde{t}$ to $\tilde{t} + \Delta_D$, and when in addition to extending the duration of the lockdown, the proportion of the population under lockdown in the neighboring municipalities at date $\tilde{t}$ is maintained constant until $\tilde{t} + \Delta_D$.

| | $\Delta_D$ | $P_{[\tilde{t}+1,\tilde{t}+\Delta_D]}$ | avg($R_{[\tilde{t}+1,\tilde{t}+21]}(*)$) | cum($I_{[\tilde{t}+1,\tilde{t}+21]}(*)$) | cum($I_{[\tilde{t}+1,\tilde{t}+21]}(*)$) |
|---|---|---|---|---|---|
| Independencia | 0 | $p^{\text{obs}}$ | 2.27 | 471 | 331 |
| Independencia | 21 | $p^{\text{obs}}$ | 1.37 [1.18, 1.55] | 136 [114, 158] | 96 [81, 111] |
| Independencia | 21 | 54.0% | 1.34 [1.16, 1.52] | 131 [110, 152] | 92 [78, 107] |
| Las Condes | 0 | $p^{\text{obs}}$ | 2.26 | 763 | 231 |
| Las Condes | 21 | $p^{\text{obs}}$ | 1.71 [1.15, 2.27] | 410 [196, 624] | 124 [59, 189] |
| Las Condes | 21 | 49.5% | 1.44 [1.26, 1.63] | 308 [275, 342] | 93 [83, 103] |
| Lo Barnechea | 0 | $p^{\text{obs}}$ | 1.83 | 364 | 293 |
| Lo Barnechea | 21 | $p^{\text{obs}}$ | 1.27 [1.21, 1.34] | 187 [176, 197] | 151 [142, 159] |
| Lo Barnechea | 21 | 53.0% | 1.19 [1.13, 1.25] | 167 [158, 176] | 135 [127, 142] |
| Ñuñoa | 0 | $p^{\text{obs}}$ | 1.71 | 422 | 168 |
| Ñuñoa | 21 | $p^{\text{obs}}$ | 1.11 [1.05, 1.17] | 212 [200, 223] | 85 [80, 89] |
| Ñuñoa | 21 | 56.5% | 1.04 [0.99, 1.10] | 191 [182, 201] | 77 [73, 80] |
| Providencia | 0 | $p^{\text{obs}}$ | 1.82 | 286 | 181 |
| Providencia | 21 | $p^{\text{obs}}$ | 1.34 [1.23, 1.46] | 193 [175, 210] | 122 [111, 133] |
| Providencia | 21 | 80.3% | 1.25 [1.14, 1.37] | 171 [155, 187] | 109 [98, 119] |
| Santiago | 0 | $p^{\text{obs}}$ | 1.95 | 2178 | 433 |
| Santiago | 21 | $p^{\text{obs}}$ | 1.23 [1.10, 1.37] | 835 [738, 933] | 166 [147, 185] |
| Santiago | 21 | 35.8% | 1.21 [1.08, 1.34] | 808 [714, 903] | 161 [142, 179] |
| Vitacura | 0 | $p^{\text{obs}}$ | 2.61 | 145 | 150 |
| Vitacura | 21 | $p^{\text{obs}}$ | 1.63 [1.55, 1.71] | 67 [63, 71] | 69 [65, 73] |
| Vitacura | 21 | 66.9% | 1.51 [1.43, 1.59] | 58 [55, 62] | 60 [57, 64] |

**Table S10: Numeric results in Figure S5C**. Observed and estimated instantaneous reproduction number and incidences when the duration of the lockdown in a given municipality is reduced from date $\tilde{t}$ to $\tilde{t} + \Delta_D$, and when in addition to reducing the duration of the lockdown, the proportion of the population under lockdown in the neighboring municipalities at date $\tilde{t}$ is maintained constant until $\tilde{t} + 7$.

| | $\Delta_D$ | $P_{(\tilde{t}+\Delta_D,\tilde{t}+7]}$ | $R_{\tilde{t}-7}(*)$ | $R_{\tilde{t}}(*)$ | $R_{\tilde{t}+7}(*)$ |
|---|---|---|---|---|---|
| Punta Arenas | 0 | $p^{\text{obs}}$ | 0.95 | 0.72 | 0.72 |
| Punta Arenas | -14 | $p^{\text{obs}}$ | 1.00 [0.88, 1.12] | 0.93 [0.76, 1.09] | 0.80 [0.60, 1.00] |
| Punta Arenas | -14 | 0.0% | 1.00 [0.88, 1.12] | 0.93 [0.76, 1.09] | 0.80 [0.60, 1.00] |
| | $\Delta_D$ | $P_{(\tilde{t}+\Delta_D,\tilde{t}+7]}$ | $I_{\tilde{t}-7}(*)$ | $I_{\tilde{t}}(*)$ | $I_{\tilde{t}+7}(*)$ |
| Punta Arenas | 0 | $p^{\text{obs}}$ | 14.91 | 10.45 | 7.97 |
| Punta Arenas | -14 | $p^{\text{obs}}$ | 14.84 [13.1, 16.58] | 13.87 [11.41, 16.32] | 11.17 [8.40, 13.93] |
| Punta Arenas | -14 | 0.0% | 14.84 [13.1, 16.58] | 13.87 [11.41, 16.32] | 11.17 [8.40, 13.93] |

**Table S11: Numeric results for the municipality of Lo Barnechea in Figure 3**. A range of hypercritical values from 0 to 1 are explored for $P_t$, letting the length of extended lockdown $\Delta_D$ vary from 0 to 14. The end of evaluation is 21 days after $\tilde{t}$. The average incidence $\text{avg}(I_{(\tilde{t},t]})$ is scaled to incidence per 100,000 people in Table S11-S14.

| Lo Barnechea | | $\text{avg}(R_{[\tilde{t}+1,t]}), t =$ | | | $\text{avg}(I_{[\tilde{t}+1,t]}), t =$ | | |
|---|---|---|---|---|---|---|---|
| $\Delta_D$ | $P_{[\tilde{t}+1,\tilde{t}+21]}$ | $\tilde{t}+7$ | $\tilde{t}+14$ | $\tilde{t}+21$ | $\tilde{t}+7$ | $\tilde{t}+14$ | $\tilde{t}+21$ |
| 0 | 0.0% | 1.49 | 1.49 | 1.34 | 6.71 | 8.29 | 8.68 |
| 0 | 25.0% | 1.49 | 1.46 | 1.33 | 6.68 | 8.02 | 8.53 |
| 0 | 50.0% | 1.48 | 1.41 | 1.30 | 6.66 | 7.65 | 8.12 |
| 0 | 75.0% | 1.48 | 1.37 | 1.29 | 6.64 | 7.31 | 7.86 |
| 0 | 100.0% | 1.47 | 1.34 | 1.31 | 6.60 | 7.13 | 7.93 |
| 7 | 0.0% | 1.29 | 1.42 | 1.47 | 5.71 | 7.35 | 9.54 |
| 7 | 25.0% | 1.29 | 1.36 | 1.39 | 5.69 | 6.93 | 8.47 |
| 7 | 50.0% | 1.28 | 1.33 | 1.32 | 5.66 | 6.69 | 7.78 |
| 7 | 75.0% | 1.28 | 1.31 | 1.28 | 5.63 | 6.56 | 7.38 |
| 7 | 100.0% | 1.27 | 1.28 | 1.24 | 5.61 | 6.37 | 6.92 |
| 14 | 0.0% | 1.29 | 1.29 | 1.38 | 5.71 | 6.46 | 8.16 |
| 14 | 25.0% | 1.29 | 1.26 | 1.32 | 5.69 | 6.28 | 7.50 |
| 14 | 50.0% | 1.28 | 1.23 | 1.27 | 5.66 | 6.11 | 6.98 |
| 14 | 75.0% | 1.28 | 1.21 | 1.23 | 5.63 | 5.94 | 6.62 |
| 14 | 100.0% | 1.27 | 1.18 | 1.19 | 5.61 | 5.79 | 6.25 |

**Table S12: Numeric results for the municipality of Providencia in Figure 3**. A range of hypercritical values from 0 to 1 are explored for $P_t$, letting the length of extended lockdown $\Delta_D$ vary from 0 to 14. The end of evaluation is 21 days after $\tilde{t}$.

| Providencia | | $\text{avg}(R_{[\tilde{t}+1,t]}), t =$ | | | $\text{avg}(I_{[\tilde{t}+1,t]}), t =$ | | |
|---|---|---|---|---|---|---|---|
| $\Delta_D$ | $P_{[\tilde{t}+1,\tilde{t}+21]}$ | $\tilde{t}+7$ | $\tilde{t}+14$ | $\tilde{t}+21$ | $\tilde{t}+7$ | $\tilde{t}+14$ | $\tilde{t}+21$ |
| 0 | 0.0% | 1.30 | 1.68 | 1.85 | 4.42 | 7.17 | 12.03 |
| 0 | 25.0% | 1.29 | 1.61 | 1.79 | 4.39 | 6.77 | 11.11 |
| 0 | 50.0% | 1.28 | 1.55 | 1.75 | 4.37 | 6.42 | 10.41 |
| 0 | 75.0% | 1.28 | 1.52 | 1.74 | 4.35 | 6.25 | 10.23 |
| 0 | 100.0% | 1.27 | 1.50 | 1.73 | 4.33 | 6.08 | 10.00 |
| 7 | 0.0% | 1.13 | 1.44 | 1.73 | 3.81 | 5.48 | 9.43 |
| 7 | 25.0% | 1.13 | 1.38 | 1.63 | 3.79 | 5.21 | 8.31 |
| 7 | 50.0% | 1.12 | 1.34 | 1.54 | 3.77 | 4.98 | 7.46 |
| 7 | 75.0% | 1.12 | 1.32 | 1.50 | 3.75 | 4.90 | 7.06 |
| 7 | 100.0% | 1.11 | 1.31 | 1.47 | 3.73 | 4.83 | 6.82 |
| 14 | 0.0% | 1.13 | 1.30 | 1.47 | 3.81 | 4.83 | 6.78 |
| 14 | 25.0% | 1.13 | 1.27 | 1.41 | 3.79 | 4.70 | 6.30 |
| 14 | 50.0% | 1.12 | 1.25 | 1.36 | 3.77 | 4.56 | 5.87 |
| 14 | 75.0% | 1.12 | 1.22 | 1.32 | 3.75 | 4.43 | 5.56 |
| 14 | 100.0% | 1.11 | 1.19 | 1.29 | 3.73 | 4.31 | 5.31 |

**Table S13: Numeric results for the municipality of Santiago in Figure 3**. A range of hyper-critical values from 0 to 1 are explored for $P_t$, letting the length of extended lockdown $\Delta_D$ vary from 0 to 14. The end of evaluation is 21 days after $\tilde{t}$.

| Santiago | | $\text{avg}(R_{[\tilde{t}+1,t]}), t =$ | | | $\text{avg}(I_{[\tilde{t}+1,t]}), t =$ | | |
|---|---|---|---|---|---|---|---|
| $\Delta_D$ | $P_{[\tilde{t}+1,\tilde{t}+21]}$ | $\tilde{t}+7$ | $\tilde{t}+14$ | $\tilde{t}+21$ | $\tilde{t}+7$ | $\tilde{t}+14$ | $\tilde{t}+21$ |
| 0 | 0.0% | 1.65 | 2.21 | 2.53 | 8.60 | 17.35 | 40.90 |
| 0 | 25.0% | 1.63 | 2.10 | 2.43 | 8.47 | 15.96 | 36.36 |
| 0 | 50.0% | 1.61 | 2.00 | 2.35 | 8.35 | 14.82 | 33.03 |
| 0 | 75.0% | 1.59 | 1.92 | 2.28 | 8.24 | 13.83 | 30.19 |
| 0 | 100.0% | 1.57 | 1.86 | 2.25 | 8.16 | 13.11 | 28.78 |
| 7 | 0.0% | 1.38 | 1.52 | 1.98 | 7.00 | 9.36 | 18.88 |
| 7 | 25.0% | 1.37 | 1.45 | 1.85 | 6.98 | 8.80 | 16.27 |
| 7 | 50.0% | 1.37 | 1.39 | 1.74 | 6.96 | 8.27 | 14.08 |
| 7 | 75.0% | 1.37 | 1.33 | 1.64 | 6.94 | 7.82 | 12.39 |
| 7 | 100.0% | 1.36 | 1.29 | 1.56 | 6.92 | 7.48 | 11.13 |
| 14 | 0.0% | 1.38 | 1.28 | 1.41 | 7.00 | 7.44 | 9.54 |
| 14 | 25.0% | 1.37 | 1.26 | 1.35 | 6.98 | 7.30 | 8.88 |
| 14 | 50.0% | 1.37 | 1.24 | 1.29 | 6.96 | 7.16 | 8.30 |
| 14 | 75.0% | 1.37 | 1.22 | 1.25 | 6.94 | 7.03 | 7.79 |
| 14 | 100.0% | 1.36 | 1.21 | 1.21 | 6.92 | 6.89 | 7.39 |

**Table S14: Numeric results for the municipality of Punta Arenas in Figure S5D**. A range of hypercritical values from 0 to 1 are explored for $P_t$, letting the length of extended lockdown $\Delta_D$ vary from -14 to 0. The end of evaluation is 21 days after $\tilde{t} - 14$.

| Punta Arenas | | avg$(R_{(\tilde{t}-14,t]}), t =$ | | | avg$(I_{(\tilde{t}-14,t]}), t =$ | | |
|---|---|---|---|---|---|---|---|
| $\Delta_D$ | $P_{(\tilde{t}+\Delta_D,\tilde{t}+7]}$ | $\tilde{t}-7$ | $\tilde{t}$ | $\tilde{t}+7$ | $\tilde{t}-7$ | $\tilde{t}$ | $\tilde{t}+7$ |
| -14 | 0.0% | 1.01 | 1.00 | 0.95 | 10.74 | 10.56 | 9.95 |
| -14 | 25.0% | 0.99 | 0.96 | 0.95 | 10.59 | 10.05 | 9.75 |
| -14 | 50.0% | 0.98 | 0.92 | 0.94 | 10.48 | 9.62 | 9.36 |
| -14 | 75.0% | 0.98 | 0.87 | 0.91 | 10.39 | 9.06 | 8.72 |
| -14 | 100.0% | 0.97 | 0.84 | 0.93 | 10.38 | 8.70 | 8.58 |
| -7 | 0.0% | 1.00 | 0.95 | 0.96 | 10.67 | 10.00 | 9.78 |
| -7 | 25.0% | 1.00 | 0.94 | 0.92 | 10.67 | 9.88 | 9.30 |
| -7 | 50.0% | 1.00 | 0.93 | 0.90 | 10.67 | 9.82 | 9.04 |
| -7 | 75.0% | 1.00 | 0.93 | 0.87 | 10.67 | 9.80 | 8.76 |
| -7 | 100.0% | 1.00 | 0.93 | 0.85 | 10.67 | 9.76 | 8.50 |
| 0 | 0.0% | 1.00 | 0.90 | 0.90 | 10.67 | 9.44 | 8.80 |
| 0 | 25.0% | 1.00 | 0.90 | 0.89 | 10.67 | 9.44 | 8.76 |
| 0 | 50.0% | 1.00 | 0.90 | 0.89 | 10.67 | 9.44 | 8.74 |
| 0 | 75.0% | 1.00 | 0.90 | 0.89 | 10.67 | 9.44 | 8.72 |
| 0 | 100.0% | 1.00 | 0.90 | 0.88 | 10.67 | 9.44 | 8.71 |
